# A Cost Analysis of 10-Year Versus 20-Year Glass Slide Retention in the Era of Whole Slide Imaging

**DOI:** 10.64898/2026.11.25.26368955

**Authors:** Matthew J. Cecchini, Rosemarie Tremblay-LeMay, David Beyer, George M. Yousef

## Abstract

**Background:** The Canadian Association of Pathologists recommends keeping glass slides for 20 years, a recommendation made before digital pathology; United States regulation requires 10 years and United Kingdom guidance permits 8 years when the block is kept. Glass is inexpensive to store, whereas digital whole slide images (WSI) incur recurring storage charges, and how often old material is requested has rarely been measured.

**Methods:** A deterministic cost model (2026 Canadian dollars) compared 10- and 20-year rules in illustrative laboratories accessioning 100,000, 500,000, and 750,000 slides/year under three strategies: glass only, glass plus WSI of every slide, and glass plus a hypothetical selective WSI archive. Requests for discarded glass were met by recuts. Because no published data describe second-decade requests, their share was set at an illustrative 0.5%, varied from 0% to 2%, and break-even thresholds were calculated. Off-site glass storage was a secondary analysis.

**Results:** For glass only, the 20-year rule cost less than the 10-year rule only if more than 0.32% of slides were requested at ages 10 to 19 years (or, at 0.5%, if recuts cost more than CAD 14.54 per slide); at 0.5% it was 6.6% to 6.9% cheaper, and at 0.01% 14.6% to 14.9% more expensive. With WSI of every slide, 20-year retention increased annual costs by 35.3% to 36.8% (CAD 153,210/year, academic laboratory). Keeping glass 20 years and WSI 10 years removed this increase. Selective WSI was cheapest only under favourable selection assumptions. Off-site glass storage after the first year added 43% to 62%.

**Conclusions:** Whether a 20-year glass rule costs more than a 10-year rule depends on how often old glass is requested, which is largely unknown; applied to digital images, a 20-year rule added substantial cost unless more than about 1.7% of slides were requested in their second decade. Flexibility in storage rules can allow laboratories to optimize storage for cost and access to diagnostic material.

## Introduction

Surgical pathology laboratories maintain archives of stained glass slides and formalin-fixed, paraffin-embedded tissue blocks to support subsequent clinical care, secondary consultations, quality assurance audits, and medicolegal inquiries. Regulatory and accreditation retention minimums were established when physical glass slides and tissue blocks constituted the sole diagnostic record, and mandated retention durations vary substantially across international jurisdictions. In the United States, the Clinical Laboratory Improvement Amendments of 1988 (CLIA) regulations require laboratories to retain histopathology slides for at least 10 years from the date of examination, a standard mirrored by the College of American Pathologists (CAP).^1,2^ In the United Kingdom, guidance from the Royal College of Pathologists and the Institute of Biomedical Science (RCPath/IBMS) permits disposal of histopathology glass slides after 8 years, provided that the tissue block is retained for 30 years and comparable sections can be recut.^3^

In Canada, the Canadian Association of Pathologists (CAP-ACP) recommends a 20-year retention period for both surgical pathology glass slides and paraffin blocks, a position formulated in 2005 prior to the clinical introduction of whole slide imaging (WSI) and reaffirmed in the 2024 British Columbia provincial guideline.^4,5^ Certain provincial statutes and community laboratory guidelines specify a 10-year minimum for adult glass slides.^6,7^ We are not aware of survey data on which period Canadian laboratories follow; in our experience, many follow the 20-year CAP-ACP recommendation. That recommendation is twice the 10-year minimum in the United States and Australia and 2.5 times the United Kingdom minimum for slides that can be replaced from the block, and requirements for retaining the corresponding digital images also differ between jurisdictions.^8–11^

During the era of conventional light microscopy, the operational cost difference between a 10-year and a 20-year slide retention policy was considered modest. Physical glass slides are relatively inexpensive to store once filed; personnel labour for initial filing, case retrieval, refiling, and end-of-retention culling accounts for much of glass archive expenditure,^12^ and few cases are recalled more than a year after diagnosis.^13^ Published data on how often glass is requested 10 or more years after diagnosis are, however, lacking. The transition to primary digital diagnosis alters this cost structure. A surgical pathology slide digitized at 40× magnification generates a compressed file of the order of 1 to 2 GB, depending on tissue area, scanner and compression, and commercial cloud archival storage incurs recurring capacity charges for every additional year of retention. As laboratories digitize high-volume surgical pathology workflows, they must determine whether legacy 20-year glass retention rules should be extended to WSI. Regulatory frameworks have begun to link digital image retention to physical slide requirements. In October 2026, the Centers for Medicare & Medicaid Services and the Centers for Disease Control and Prevention published a proposed rule under CLIA that would amend 42 CFR §493.1105(a)(7)(i)(B) to require retention of both the histopathology glass slide and, if applicable, the corresponding digital slide image for 10 years; the rule had not been finalized at the time of writing.^8^ In Canada, statutory retention schedules do not explicitly address diagnostic WSI, and the CAP-ACP telepathology guideline defers image retention policy to regional jurisdictions.^11^ Across Europe and North America, clinical practice remains heterogeneous: some fully digital laboratories retain all diagnostic WSI indefinitely or for the full statutory period, whereas others purge WSI after several months or selectively archive cases of high clinical or educational value.^13–15^

Implementation reports of digital pathology frequently identify digital storage as a major expense,^14,15^ but in the archive and implementation literature we reviewed we did not identify a study that isolates the cost of the retention period itself. We therefore developed an activity-based cost model to evaluate the annual and longitudinal costs of 10-year versus 20-year retention policies in illustrative small community, academic, and large academic pathology laboratories operating glass-only, universal WSI, and selective WSI archives, and compared on-site with commercial off-site storage of glass slides. Because several key inputs have not been measured, we present the results as scenario and threshold analyses rather than as estimates for any particular laboratory.

## Methods

### Study Design and Reporting Standards

We constructed a deterministic activity-based cost model from the perspective of the anatomic pathology laboratory, capturing the capital, space, vendor, and personnel costs required to store, retrieve, recut, and purge archived diagnostic material. Costs are economic costs: they include allocated staff time, annualized cabinet capital, and the opportunity cost of floor space, which are not all cash expenditures that change immediately with a retention policy. The study was designed and reported in accordance with the Consolidated Health Economic Evaluation Reporting Standards 2022 (CHEERS 2022) statement^16^ (Supplementary File 2). All monetary values are expressed in 2026 Canadian dollars (CAD); prices quoted in United States dollars (USD) were converted at the Bank of Canada daily exchange rate of 1.4257 CAD per USD^17^ without further inflation adjustment. The spreadsheet model with live formulas is provided in Supplementary File 1, and all input parameters, data sources, and sensitivity ranges are detailed in Supplementary File 3 (Supplementary Tables S1 to S11). Each input is classified in Supplementary Table S1 as a published value, a published value used as a proxy, or an assumption; the request rates, the second-decade request share, the selection rule, labour times, and several cabinet inputs are assumptions because published data are lacking.

### Laboratory Tiers, Archival Scenarios, and Retention Policies

To reflect a range of Canadian practice, we modelled three illustrative laboratory volume tiers: a small community hospital laboratory accessioning 100,000 slides per year, an academic medical centre laboratory accessioning 500,000 slides per year, and a large academic health network accessioning 750,000 slides per year, assuming a mean of 3 slides per surgical pathology case. The same value was used for slides per requested case. Each laboratory tier was evaluated across three archival scenarios under both a 10-year and a 20-year retention rule:

1. Glass-only archive. Physical glass slides are retained on site for 10 or 20 years. Paraffin tissue blocks are retained for 20 years under both rules; consequently, any clinical request for a case aged 10 to 19 years under the 10-year slide rule is fulfilled by pulling the retained tissue block, recutting serial sections, and performing hematoxylin and eosin (H&E) staining.
2. Glass plus universal WSI archive. Every glass slide is digitized at 40× magnification for primary diagnosis, and both the physical glass slide and its corresponding WSI are retained for the same mandated duration (10 or 20 years).
3. Glass plus selective WSI archive. Physical glass slides are retained for 10 or 20 years as in Scenario 1, but long-term cloud archival of WSI is restricted to a curated subset of 10% of slides flagged at sign-out (such as complex oncology, consultation, teaching, or medicolegal cases), which is assumed in the base case to capture 90% of digitally servable retrieval requests. This selection performance is hypothetical; no validated method has been shown to achieve it, and the effort of selection was not costed.

Because slide scanning, image management system (IMS) licensing, local area network infrastructure, and short-term hot-tier caching during active primary sign-out are incurred irrespective of long-term archival duration, these upstream operational costs were excluded. In secondary analyses, we evaluated the United Kingdom 8-year retention rule and hybrid medium-specific policies in which physical glass slides and WSI are retained for different durations (10 versus 20 years).

### Clinical Retrieval Demand and Block Recutting

Retrieval demand is the least well documented input. Published institutional series report roughly 2.7 to 12 glass requests per 100 accessioned cases per year,^18,19^ and digital access logs show that most first accesses of an image occur within months of scanning,^20^ but we found no study reporting requests by specimen age over two decades. We therefore assumed that each year’s cases generate 5.0 retrieval requests per 100 accessioned cases while aged 0 to 9 years, where a request is one case (3 slides) and the rate is expressed per 100 cases accessioned per year rather than as an annual probability for every stored case. Of these requests, 85% were assumed to occur within the first 12 months after sign-out and the remaining 15% were spread evenly across years 1 through 9 (0.083% of cases per year at each age). For cases aged 10 to 19 years, a small fraction of slides are requested for late clinical review, recurrence comparison, or medicolegal inquiry. Under a 20-year glass retention policy, these late requests are satisfied by retrieving the archived glass slides. Under a 10-year glass retention policy, the original glass slides have been culled, requiring retrieval of the paraffin block, microtomy recutting, and H&E staining of 3 slides per case (total cost, CAD 79.97 per case request). The share of slides requested at ages 10 to 19 years is unknown. Ali et al. reported that 390 of 162,761 cases (0.24%) reported in a UK department between July 2011 and December 2015 were recalled from off-site storage more than 12 months after diagnosis;^13^ this proportion is not an age-specific rate and does not describe the second decade. Radiology data suggest that old images are opened more often when they are readily available.^22^ We therefore used 0.5% of an annual slide cohort as an illustrative base value, tested values from 0% to 2%, and report the share at which the 20-year and 10-year rules cost the same. The model assumes that a recut substitutes for the original slide; it does not capture exhausted tissue or recuts that fail to reproduce the original diagnostic material. No retrieval requests were modelled at 20 years or beyond, when paraffin blocks are also eligible for disposal. Whenever a retained WSI was available in the digital archive, retrieval requests were fulfilled electronically, except for a 10% residual fraction of requests requiring physical glass slides for specialized microscopy (such as polarization), recuts, or external slide loan.^18,23^

### Resource Valuation and Unit Costs

Laboratory technical labour was valued at CAD 0.74 per productive minute, derived from the median hourly wage for a medical laboratory assistant (NOC 33101) in an Ontario economic region (CAD 28.14)^24^ inflated to 2026 with the Statistics Canada consumer price index, with an assumed 27% benefits load and an assumed 85% productive time fraction. On-site glass slide filing (0.25 minutes/slide) and end-of-retention culling (0.05 minutes/slide) were modelled as one-time per-slide activities invariant to retention duration at steady state. Each physical glass retrieval event was assumed to require 15 minutes of staff labour (locating, pulling, logging, and refiling 3 slides) plus a 0.02% expected slide-loss replacement allowance, yielding CAD 11.11 per glass request (Supplementary Table S1); these labour times are assumptions, as we found no time-motion data. Unstained recutting and H&E staining were valued at USD 16.10 (CAD 22.95) per slide from a US academic core laboratory’s internal fee schedule,^25^ used as a proxy because we did not identify a published Canadian cost; the marginal in-house cost may be lower or higher. On-site glass archives were housed in modular 12-unit steel slide cabinet stacks (51,000 slides per stack at an assumed 85% usable capacity),^26^ with capital costs annualized over an assumed 20-year cabinet lifespan; the cabinet price is the list price shown on the cited product page on November 25, 2026 (USD 199 per cabinet), less an assumed 10% volume discount; the price of the cabinet base and the discount are assumptions. Floor space was valued at commercial gross office rent, as a proxy for hospital space, including aisle and circulation allowances.^27^

Digital WSI files (1.5 GB per slide at 40× magnification, an assumed value within published ranges^20,23^) were archived in Microsoft Azure Archive Blob Storage (Locally Redundant Storage, Canada Central region) under a 3-year reserved capacity commitment (100 TiB and 1 PiB reservation blocks, with file sizes treated as GiB when sizing reservations), the lowest-priced Canadian-region archive option among those evaluated at published list prices.^28^ On a continuous per-slide-year basis, cloud archival storage of a 1.5 GB WSI cost CAD 0.0360 per year, or 5.0 times the on-site physical storage cost of a glass slide (CAD 0.0073 per slide-year). Each digitally fulfilled retrieval request triggered asynchronous rehydration of the 3-slide case (4.5 GB) from the archive tier, which takes up to 15 hours at standard priority,^29^ retention of the restored copy in hot-tier storage for 6 months, and one internet egress download. Images in active use for primary diagnosis are held in a separate diagnostic cache whose cost does not depend on the retention rule and was excluded. Alternative Canadian cloud archive tiers (Amazon Web Services S3 Glacier Deep Archive, Azure Archive pay-as-you-go, and Google Cloud Archive) were evaluated as scenarios.^30,31^ The model keeps images in Canadian cloud regions; we did not assess contractual, availability, recovery, privacy, or record-preservation requirements beyond data location.

### Outcome Measures and Sensitivity Analyses

The primary outcome measure was the modelled steady-state annual cost (CAD/year) of operating a fully populated archive under each retention rule. The difference between the 20-year and 10-year rules represents the recurring annual premium (or saving) of maintaining the second decade of archived material. As a secondary outcome, we calculated the 30-year net present cost (NPC) starting from an empty archive, discounted at the 1.5% real annual reference-case rate recommended by Canada’s Drug Agency (CDA-AMC; sensitivity range, 0% to 3%);^32^ in this calculation cabinets were charged as an equivalent annual cost while in use, consistent with the steady-state model, and reserved cloud capacity was charged at one third of each 3-year commitment per year. Deterministic one-way sensitivity analyses were performed for the second-decade request share (0% to 2%), selective WSI capture, overall retrieval request rates, residual glass pull rates, retrieval labour time, hot-tier retention duration (3 to 12 months), network egress pathway, and discount rate (Supplementary Tables S5 and S6). Cloud provider tier, 20× scanning (0.5 GB/slide), and off-site glass storage change several linked costs and are reported as scenarios. We calculated two-way break-even thresholds for the second-decade request share against recut cost, and for selective WSI coverage against the selection effort that would offset its saving (Supplementary Figure S3 and Supplementary Table S11). We did not undertake a probabilistic analysis, because the inputs that matter most lack data from which to specify distributions.

### Secondary Analysis: Off-Site Glass Storage

Because hospital space is constrained, we compared on-site storage of glass slides with commercial off-site storage. We did not identify a public Canadian tariff for slide storage, so we used the national price list that Iron Mountain submitted to a United States public-sector purchasing cooperative in 2025, converted at the same exchange rate without inflation adjustment.^33^ Two contracting models were priced. Under the pathology lines, each case is received, listed, retrieved, and refiled individually (temperature-controlled storage USD 2.55 per cubic foot per month; receiving and listing USD 7.27 per case; retrieval and refiling USD 7.02 per case), giving an intake cost of CAD 3.67 per slide. Under the climate-controlled records lines, slides are sent in cartons indexed by the laboratory (storage USD 2.15 per cubic foot per month; receiving USD 1.97 per cubic foot, the general records line, which differs immaterially from the climate-controlled line of USD 1.90; retrieval and refiling of a 1.2 cubic foot carton USD 8.45), giving an intake cost of CAD 0.21 per slide and a storage cost of CAD 0.0130 per slide-year. Whether these terms apply to pathology material in Canada, and the packing density and courier batching assumed, would need confirmation with a vendor. In the hybrid policy, slides were filed on site for their first year, when most first-decade requests (85%) occur, then packed (0.25 minutes per slide) and transferred to the vendor; requests for older cases were delivered the next day (an assumed 20 requests per courier trip and 6 minutes of in-house labour per request; CAD 24.30 per request under the records lines), and the vendor destroyed slides at the end of retention. We also evaluated transfer to the vendor at accession and valued on-site floor space at new hospital construction cost (CAD 610 per m² per year). Paraffin blocks remained on site in all policies, so recuts always incurred the on-site block retrieval cost.

## Results

### Scenario 1: Glass-Only Archive

Under a 10-year retention policy, the modelled steady-state annual cost of a glass-only archive was CAD 61,454 for the small community laboratory, CAD 306,897 for the academic laboratory, and CAD 460,160 for the large academic laboratory, corresponding to CAD 0.61 per accessioned slide (Table 1). At the illustrative base share of 0.5%, extending glass slide retention from 10 to 20 years reduced steady-state annual costs by 6.6% to 6.9% across the three laboratory tiers (Figure 1), saving CAD 4,060 per year in the small laboratory, CAD 21,042 per year in the academic laboratory, and CAD 31,562 per year in the large laboratory (CAD 0.57 versus 0.61 per accessioned slide). Under the 10-year rule, the assumed second-decade requests for discarded glass generated 167, 833, and 1,250 case recut requests per year, costing CAD 13,329, CAD 66,643, and CAD 99,964 annually, whereas the cabinets and floor space for a second decade of glass cost CAD 7,417, CAD 36,343, and CAD 54,514 per year. The direction of this result depends on the assumed request share (see Sensitivity Analyses). Under the 10-year rule, personnel labour for initial slide filing and end-of-retention culling accounted for 36% of total annual cost, retrieval labour for 30%, block recuts for 22%, and physical storage (cabinet amortization and floor space) for only 12% (Figure 2).

**Figure 1.**
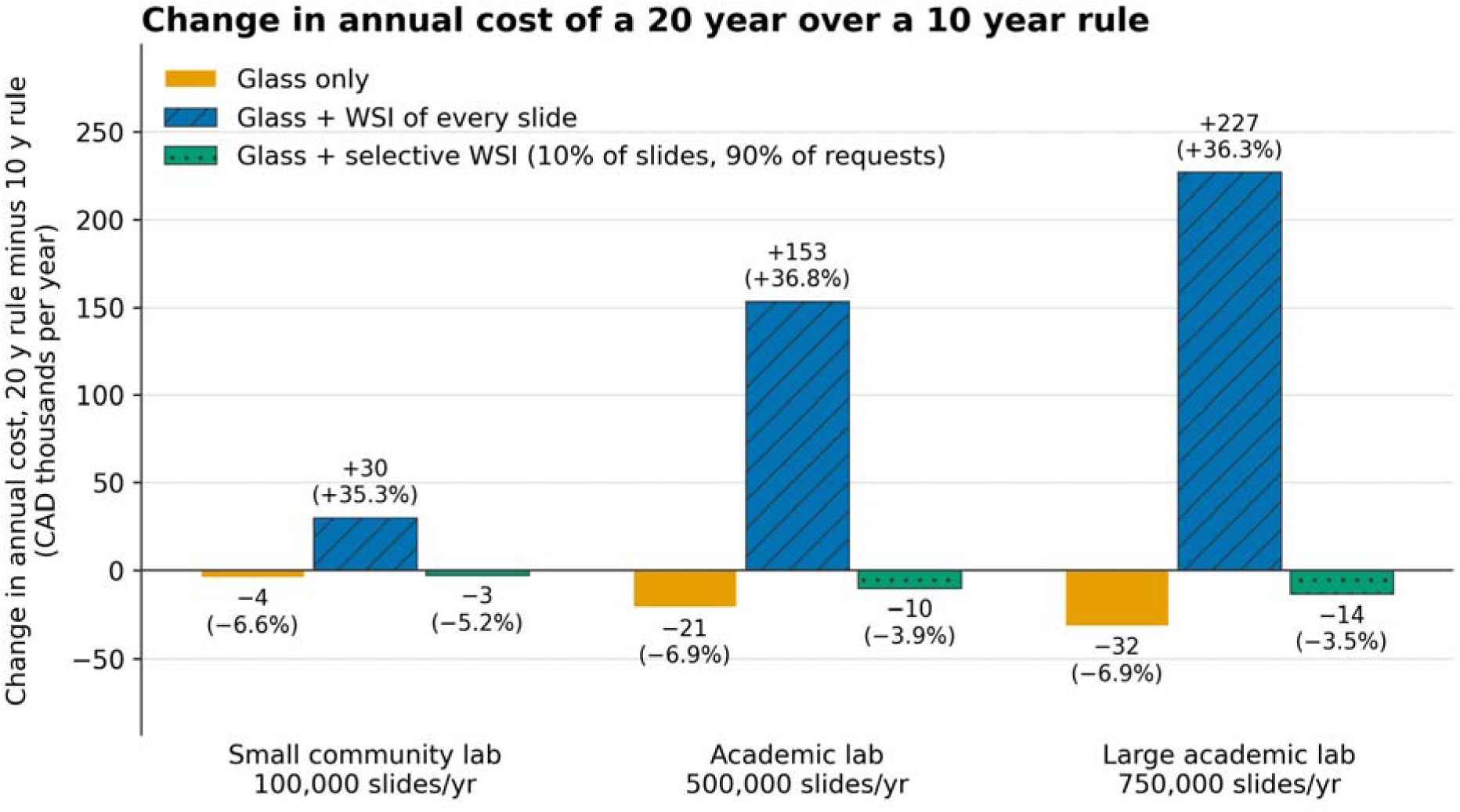
Change in annual cost of a 20-year over a 10-year retention rule. Change in steady-state annual cost of a 20-year over a 10-year rule (glass and images kept for the same period) for the three laboratories (100,000, 500,000 and 750,000 slides per year) and three scenarios. Labels give the change in CAD thousands per year and, in brackets, as a percentage of the 10-year cost: glass only −4 / −21 / −32 (−6.6% / −6.9% / −6.9%); glass plus WSI of every slide +30 / +153 / +227 (+35.3% / +36.8% / +36.3%); glass plus selective WSI −3 / −10 / −14 (−5.2% / −3.9% / −3.5%) (small / academic / large). Under a 10-year rule, requests for discarded glass (an illustrative 0.5% of slides, at ages 10 to 19 years) are met by recutting the block, which costs more than keeping the glass for a second decade; the glass-only result reverses below a share of about 0.32%, or below a recut cost of about CAD 15 per slide (Supplementary Figure S3). CAD, Canadian dollars (2026); WSI, whole slide images.

**Figure 2.**
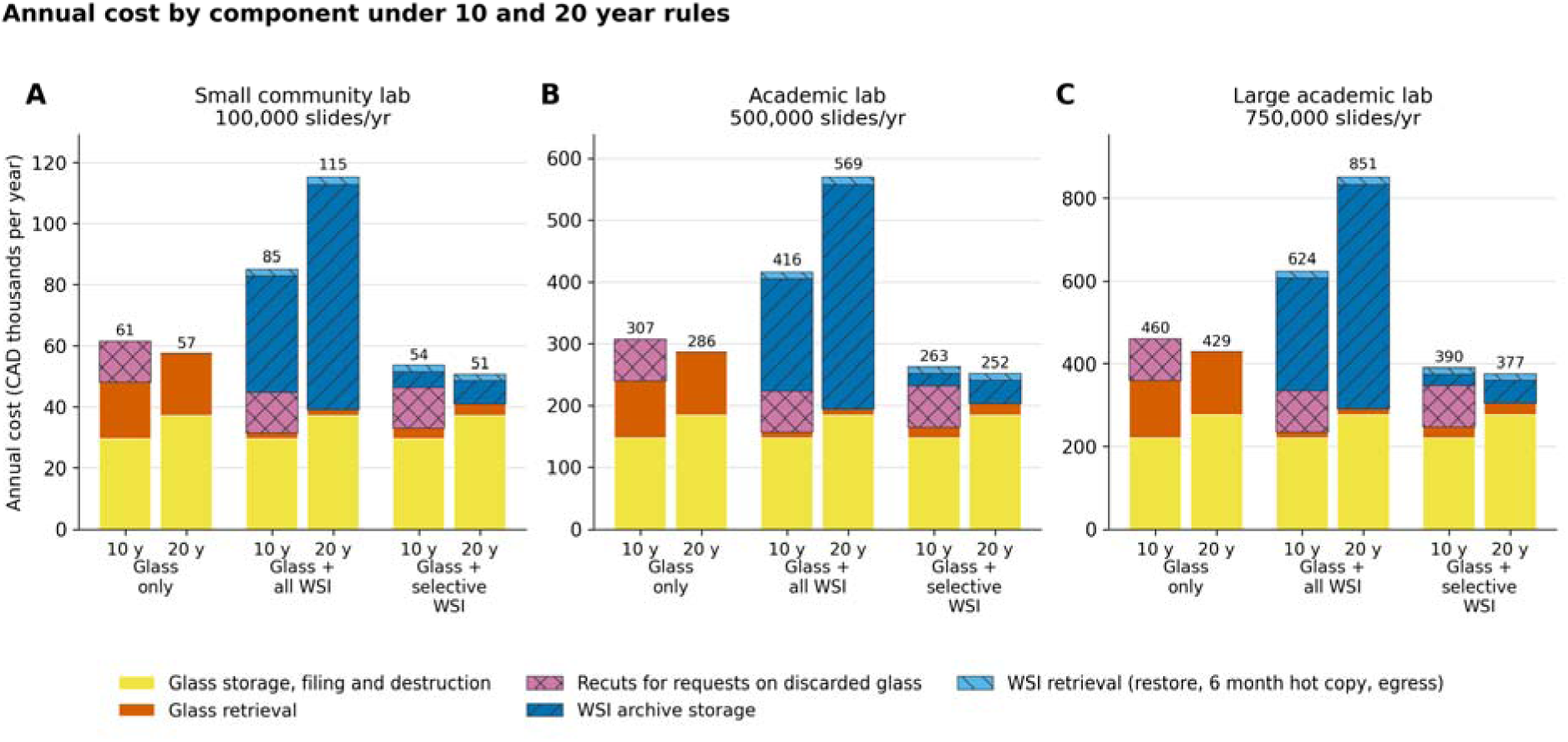
Steady-state annual cost of each scenario under a 10 and a 20-year rule, by cost component, for (A) the small community, (B) the academic and (C) the large academic laboratory; y axes differ between panels. The number above each bar is the total in CAD thousands per year. Glass retrieval is the labour of pulling and refiling requested slides. Recuts are block pulls, recuts and H&E staining for requests on glass discarded at 10 years (CAD 13,329 / 66,643 / 99,964 per year under a 10-year rule). WSI retrieval is the cost of restoring requested cases from the archive tier (Azure Archive, Canada Central), keeping them 6 months in hot storage and downloading them once. Selective WSI keeps images of 10% of slides, assumed to cover 90% of requests. Scanning, image management software, networks and the diagnostic image cache are excluded. CAD, Canadian dollars (2026); WSI, whole slide images; y, years.

**Table 1.** Steady-State Annual Storage and Retrieval Cost by Laboratory, Scenario and Retention Rule.

| **Laboratory** | **Scenario** | **10 y rule (CAD/y)** | **20 y rule (CAD/y)** | **Change with 20 y rule (CAD/y)** | **Change** | **Per slide, 10 y / 20 y (CAD)** |
|---|---|---|---|---|---|---|
| Small community | Glass only | 61,454 | 57,394 | –4,060 | –6.6% | 0.61 / 0.57 |
|  | Glass + WSI of every slide | 85,114 | 115,138 | +30,024 | +35.3% | 0.85 / 1.15 |
|  | Glass + selective WSI | 53,614 | 50,838 | –2,775 | –5.2% | 0.54 / 0.51 |
| Academic | Glass only | 306,897 | 285,856 | –21,042 | –6.9% | 0.61 / 0.57 |
|  | Glass + WSI of every slide | 416,098 | 569,308 | +153,210 | +36.8% | 0.83 / 1.14 |
|  | Glass + selective WSI | 262,526 | 252,409 | –10,118 | –3.9% | 0.53 / 0.50 |
| Large academic | Glass only | 460,160 | 428,598 | –31,562 | –6.9% | 0.61 / 0.57 |
|  | Glass + WSI of every slide | 624,298 | 851,191 | +226,893 | +36.3% | 0.83 / 1.13 |
|  | Glass + selective WSI | 390,348 | 376,799 | –13,548 | –3.5% | 0.52 / 0.50 |

Abbreviations: CAD, Canadian dollars (2026); WSI, whole slide images; y, years. Illustrative laboratories producing 100,000, 500,000 and 750,000 slides per year. Modelled steady-state annual economic cost with a full archive; glass and images are kept for the same period. Totals include glass storage, filing and destruction, glass retrieval, recuts of glass discarded at 10 years (0.5% of slides requested at ages 10 to 19 years, an illustrative assumption with no published data; 0% to 2% and break-even in Supplementary Tables S6 and S11), image archive storage and digital retrieval (archive restore, 6-month hot copy and egress). Negative values: the 20-year rule costs less. Selective WSI: 10% of slides hypothetically covering 90% of requests that an image can serve. Per slide: annual cost divided by slides accessioned per year. Components: Figure 2 and Supplementary Table S4.

### Scenario 2: Glass Plus Universal WSI Archive

Retaining a 40× WSI of every slide alongside the physical glass archive reduced manual glass pulls by 90% but increased total archive expenditure at every laboratory scale (Table 1). Under a 10-year retention rule, universal WSI plus glass cost CAD 85,114, CAD 416,098, and CAD 624,298 per year across the small, academic, and large laboratories (CAD 0.83 to 0.85 per accessioned slide), representing a 35.6% to 38.5% increase over glass-only retention. Under a 20-year retention rule, total annual expenditure rose to CAD 115,138, CAD 569,308, and CAD 851,191 per year (CAD 1.13 to 1.15 per slide). Thus, whereas at the base request share a 20-year rule reduced the cost of a glass-only archive, extending retention from 10 to 20 years when universal WSI were kept increased annual archive costs by 35.3% to 36.8% (CAD +30,024, +153,210, and +226,893 per year) (Figure 1). Under the 10-year rule, cloud storage capacity charges accounted for 94.3% to 94.5% of total digital cost (96.7% to 96.8% under the 20-year rule), whereas digital retrieval (archive rehydration fees, 6-month hot-tier caching, and data egress) represented 5.5% to 5.7% (Figure 2). Per slide, glass incurred filing at accession (CAD 0.18) and destruction at disposal (CAD 0.04) and then CAD 0.0073 per slide-year for cabinets and space, whereas a 40× WSI incurred CAD 0.036 per slide-year, so the cumulative storage cost of an image exceeded that of its glass slide after 7.7 years (Figure 3). Maintaining both glass and universal WSI for 20 years cost 1.85 to 1.87 times as much annually as maintaining a 10-year glass-only archive.

**Figure 3.**
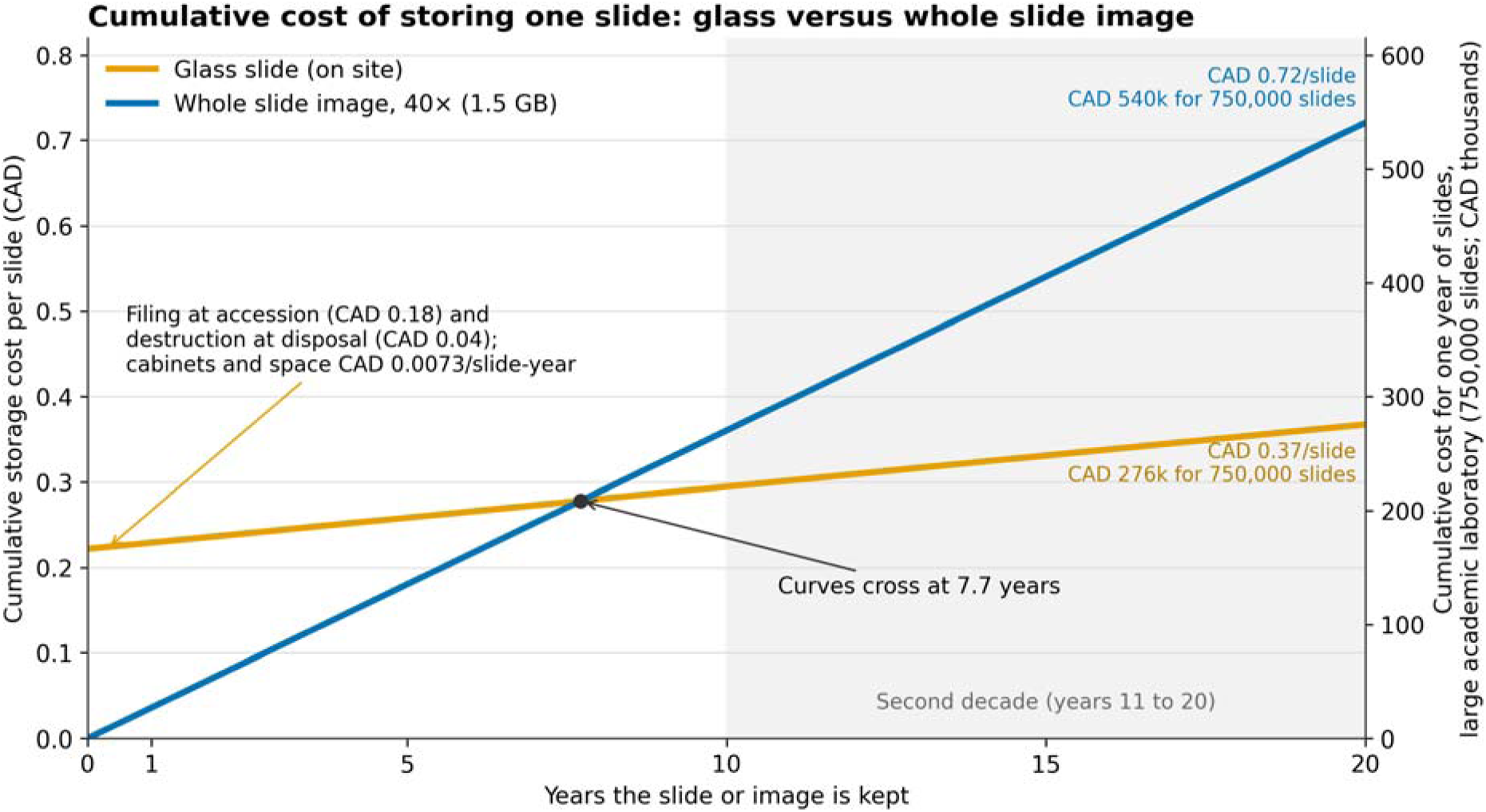
Cumulative storage cost of one slide kept as glass or as a whole slide image, by years kept. Each curve is the undiscounted cost of keeping one slide for the number of years shown. Glass kept on site incurs filing at accession (CAD 0.18) and destruction at disposal (CAD 0.04), and cabinets and floor space of CAD 0.0073 per slide-year (cabinet capital annualized while in use, as in the main model). A 40× WSI (1.5 GB) costs CAD 0.036 per slide-year in the base archive tier (Azure Archive, 3-year reserved capacity, Canada Central), 5.0 times the glass rate. The curves cross at 7.7 years. The right axis gives the same cumulative cost for one year of slides in the large academic laboratory (750,000 slides): at 20 years, CAD 0.37 per slide (CAD 276k) for glass versus CAD 0.72 per slide (CAD 540k) for a 40× WSI; this is the cost of one annual cohort, not a recurring annual budget. Storage and handling only: retrieval, recuts and digital restore costs (Figures 1 and 2) are excluded. In the WSI scenarios the glass is kept as well, so the curves compare storage media rather than scenarios. CAD, Canadian dollars (2026); WSI, whole slide images.

### Scenario 3: Glass Plus Selective WSI Archive

Archiving WSI for a curated 10% subset of slides assumed to capture 90% of digitally servable requests was the least expensive strategy in the base case across all laboratory sizes and retention durations (Table 1), under the hypothetical selection assumption. Under a 10-year rule, selective WSI plus glass cost CAD 53,614, CAD 262,526, and CAD 390,348 per year (CAD 0.52 to 0.54 per slide), yielding net annual savings of 12.8% to 15.2% (CAD 7,840 to 69,813 per year) compared with a 10-year glass-only archive. These savings occurred because a compact cloud footprint (150 to 1,125 TB at 10 years) reduced physical glass pulls within the image retention period by 81.0%. Under a 20-year rule, selective WSI cost 11.4% to 12.1% less than a 20-year glass-only archive and 3.5% to 5.2% less than selective WSI under a 10-year rule, because retained glass served second-decade requests without recuts (CAD 252,409 versus CAD 262,526 per year in the academic laboratory).

### Decoupled Media Retention Policies and Jurisdiction Comparisons

When every slide is digitized for primary diagnosis, decoupling the WSI retention period from the physical glass retention period eliminated the cost penalty of a 20-year glass rule (Table 2). In the academic laboratory, retaining physical glass slides for 20 years to satisfy CAP-ACP guidance while purging WSI after 10 years reduced annual expenditure from CAD 569,308 (both media kept 20 years) to CAD 395,057 per year, saving CAD 174,251 annually, and cost CAD 21,042 less than keeping both media for 10 years, because the retained glass served second-decade requests without recuts. Conversely, retaining glass slides for 10 years and selective WSI for 20 years cost CAD 226,969 per year in the academic laboratory, below the cost of either glass-only archive. Applying an 8-year glass rule within this Canadian model (blocks kept 20 years; the United Kingdom provisions for blocks and irreplaceable slides were not modelled) showed that shortening glass retention below 10 years did not reduce costs for a glass-only archive (+3.8% versus 10 years; Supplementary Table S7 and Supplementary Figure S1), because cases aged 8 and 9 years still generate sufficient retrieval demand that block recutting expenses outweigh the 2-year reduction in cabinet storage. The 8-year rule was less expensive than a 10-year rule only when universal WSI were retained (−5.0% to −5.8%).

**Table 2.** Separate Retention Periods for Glass and Images: Steady-State Annual Cost (CAD per Year)

| **Policy** | **Glass kept (y)** | **Images kept (y)** | **Small community** | **Academic** | **Large academic** |
|---|---|---|---|---|---|
| Glass only, 20-year rule | 20 | None | 57,394 | 285,856 | 428,598 |
| Glass only, 10-year rule | 10 | None | 61,454 | 306,897 | 460,160 |
| Glass + WSI of every slide, same period | 20 | 20 | 115,138 | 569,308 | 851,191 |
| Glass + WSI of every slide, shorter for images | 20 | 10 | 81,054 | 395,057 | 592,735 |
| Glass + WSI of every slide, shorter for glass | 10 | 20 | 108,868 | 538,704 | 805,284 |
| Glass + selective WSI, same period | 20 | 20 | 50,838 | 252,409 | 376,799 |
| Glass + selective WSI, shorter for images | 20 | 10 | 49,554 | 241,485 | 358,785 |
| Glass + selective WSI, shorter for glass | 10 | 20 | 45,602 | 226,969 | 338,640 |

Abbreviations: CAD, Canadian dollars (2026); WSI, whole slide images; y, years. Blocks are kept 20 years in every policy, so requests for discarded glass are met by a recut unless an image is kept. Selective WSI: 10% of slides assumed to cover 90% of requests (optimistic; see Supplementary Figure S2). Same inputs as Table 1; Supplementary Table S8.

### Sensitivity Analyses

The economic advantage of selective WSI archival was highly sensitive to how well cases are selected at sign-out (Supplementary Table S5 and Supplementary Figure S2). When the 10% archived subset followed an illustrative imperfect capture curve (exponent g = 3, capturing 27.1% of requests), selective WSI approximately broke even with glass-only retention (CAD +1,254, +1,097, and −1,611 per year across the three laboratories). Under random 10% selection, selective WSI was more expensive than glass-only retention by 3.7% to 6.1%. A 10% selective archive must capture at least 26% to 36% of digitally servable requests under a 10-year rule (and 47% to 49% under a 20-year rule) to offset its cloud storage and retrieval costs. Selection effort was not costed: the base-case saving would be cancelled by selection effort costing CAD 0.24 to 0.28 per case accessioned, about 19 to 23 seconds of laboratory assistant time (Supplementary Figure S3B).

The direction of the glass-only result depended on the second-decade request share and on the cost of a recut (Supplementary Table S6, Supplementary Table S11 and Supplementary Figure S3A). The 20-year and 10-year glass rules cost the same when the share of slides requested at ages 10 to 19 years multiplied by the recut and staining cost per slide equalled the cost of storing a slide for a second decade (CAD 0.073 per slide in the academic laboratory). At CAD 22.95 per recut slide, a 20-year glass rule cost more than a 10-year rule when fewer than about 0.32% of slides were requested at ages 10 to 19 years (0.032% of cases per year); at a 0.5% share it cost more if recut and staining cost less than CAD 14.54 per slide. At the previously assumed share of 0.01%, a 20-year rule increased glass-only costs by 14.6% to 14.9%, and at 0.1% by 9.8% to 10.1%, whereas at 1% and 2% it reduced them by 20.8% to 21.0% and 37.9% to 38.1%. For universal WSI, a 20-year rule increased costs at every request share up to approximately 1.7% (by 17.4% to 62.3% across the 0% to 1% range) and reduced them only at 2% (−6.6% to −7.0%).

Across the other one-way analyses and scenarios, the glass-only 20-year rule remained within −8.1% to +1.3% of the 10-year rule at the base request share, whereas the universal WSI increment ranged from +8.6% to +52.5% (Supplementary Table S6). Substituting alternative Canadian cloud archive tiers increased the 20-year universal WSI increment in the academic laboratory from +36.8% (Azure reserved) to +43.5% (AWS Glacier Deep Archive), +43.5% (Azure pay-as-you-go), and +52.4% (Google Cloud Archive); these tiers differ in retrieval times and service terms, so the comparison is of list prices only. A 20× scanning scenario (0.5 GB/slide) reduced the 20-year universal WSI cost increment to 10.9% to 11.3% and made a 10-year universal WSI archive less expensive than a 10-year glass-only archive in the academic laboratory (CAD −18,908/year); 20× images are not interchangeable with 40× images for every diagnostic purpose. Over a 30-year horizon from an empty archive (discounted at 1.5%/year; Supplementary Table S9), the net present cost of a 20-year over a 10-year rule in the academic laboratory changed by −6.2% (CAD −387,870) for glass only versus +19.4% (CAD +1,578,199) for universal WSI.

### Off-Site Glass Storage (Secondary Analysis)

Commercial off-site storage increased costs in every laboratory, scenario, and retention rule (Table 3). Keeping glass on site for its first year and then transferring it to the vendor under the records lines increased glass-only steady-state costs by 43% to 45% under a 10-year rule and by 60% to 62% under a 20-year rule (CAD 458,234 versus CAD 285,856 per year in the academic laboratory). Vendor storage cost 1.8 times as much per slide-year as on-site cabinets, each slide was handled twice (filing, then packing for transfer), and each off-site request cost CAD 24.30 versus CAD 11.11 on site. Under the pathology lines, per-case receiving and listing fees raised glass-only costs 7.1- to 7.7-fold. Sending glass to the vendor at accession rather than after 1 year changed glass-only costs by less than 1%. Valuing on-site floor space at new hospital construction cost raised on-site glass-only costs by only 5% to 11%; on-site storage would cost as much as the hybrid off-site policy only if floor space cost approximately CAD 2,027 per m² per year (glass only, 20-year rule, academic laboratory), 7.2 times the office rent used in the base case. For universal WSI, in which 90% of requests are served digitally, vendor storage from accession added 9% to 13% and the hybrid policy 26% to 29%. With glass off site, a 20-year glass-only rule cost 4.2% more than a 10-year rule at the base request share, because off-site storage of the second decade costs more than on-site cabinets.

**Table 3.** Glass-Only Archive Kept On Site or With a Commercial Off-Site Vendor: Steady-State Annual Cost (CAD per Year)

| **Storage of glass slides** | **Glass kept (y)** | **Small community** | **Academic** | **Large academic** | **Academic, vs on site** |
|---|---|---|---|---|---|
| On site (base case) | 10 | 61,454 | 306,897 | 460,160 | Reference |
| On site (base case) | 20 | 57,394 | 285,856 | 428,598 | Reference |
| On site, floor space at new hospital build cost | 10 | 64,748 | 323,207 | 484,542 | +5.3% |
| On site, floor space at new hospital build cost | 20 | 63,983 | 318,310 | 477,197 | +11.4% |
| 1 y on site, then vendor (records lines) | 10 | 89,269 | 439,715 | 658,744 | +43.3% |
| 1 y on site, then vendor (records lines) | 20 | 92,972 | 458,234 | 686,522 | +60.3% |
| 1 y on site, then vendor (pathology lines) | 10 | 436,536 | 2,176,051 | 3,263,247 | +609.0% |
| 1 y on site, then vendor (pathology lines) | 20 | 442,316 | 2,204,949 | 3,306,595 | +671.4% |
| Vendor from accession (records lines) | 10 | 90,018 | 443,463 | 664,365 | +44.5% |
| Vendor from accession (records lines) | 20 | 93,722 | 461,981 | 692,143 | +61.6% |

Abbreviations: CAD, Canadian dollars (2026); y, years. Glass only; paraffin blocks stay on site for 20 years in every option, and requests for glass discarded at 10 years are met by a recut. Vendor prices: Iron Mountain public national price list (2025; USD converted at 1.4257).<sup>33</sup> Records lines: slides sent in cartons indexed by the laboratory (intake CAD 0.21 per slide, storage CAD 0.0130 per slide-year, CAD 24.30 per off-site request). Pathology lines: each case received, listed and retrieved individually (intake CAD 3.67 per slide). Hybrid: slides kept on site for their first year, when most requests occur, then sent to the vendor. Vendor terms, packing density and courier batching would need confirmation for Canadian pathology material. New hospital build: on-site floor space valued at CAD 610 instead of CAD 282 per m² per year. Glass plus WSI of every slide and glass plus selective WSI: Supplementary Table S10 and Supplementary File 1, sheet OffSite.

## Discussion

This analysis shows a divergence between the cost dynamics of physical glass archives and digital WSI archives. In a glass-only workflow, whether extending slide retention from 10 to 20 years adds or saves money depends on two inputs that have not been measured: how often glass is requested in its second decade and what a recut costs. A second decade of cabinet storage cost about CAD 0.073 per slide, so a 20-year rule saved money when the request share multiplied by the recut cost per slide exceeded that amount; at CAD 22.95 per recut slide the break-even share was about 0.32%, and no published data establish which side of this threshold Canadian laboratories are on. Physical glass slides are inexpensive to house once accessioned: high-density steel cabinets occupy a modest footprint, representing only 12% of annual glass archive costs at 10 years, while the larger labour costs of initial filing, culling, and early post-diagnostic retrieval do not depend on whether slides are discarded at year 10 or year 20. Digital pathology reverses this relationship. At 40× magnification, storing a 1.5 GB WSI in the lowest-priced Canadian archive tier evaluated costs 5.0 times as much per slide-year as on-site cabinet storage of the physical slide. Because the cost of glass is incurred mostly at filing and disposal, whereas the cost of a WSI accrues every year it is kept, the cumulative storage cost of a 40× image overtakes that of its glass slide at 7.7 years (Figure 3). Consequently, when laboratories archive a WSI of every slide, extending retention from 10 to 20 years increased total annual archive cost by 35% to 37% (CAD 153,210 per year in a 500,000-slide academic laboratory) at the base request share, and the increase persisted at request shares up to about 1.7%.

These findings are relevant to Canadian anatomic pathology laboratories as they transition to digital sign-out. In Canada, the CAP-ACP published national guidance in 2005, prior to the clinical adoption of WSI, recommending that surgical pathology glass slides and tissue blocks be retained for 20 years, a timeframe reaffirmed in 2024 British Columbia guidance.^4,5^ Provincial hospital regulations and community laboratory guidelines specify a 10-year minimum for glass slides,^6,7^ and we are not aware of data on which period laboratories follow. The CAP-ACP recommendation is twice the 10-year minimum in the United States and Australia and 2.5 times the 8-year minimum for replaceable slides in the United Kingdom.^1–3,9^ A 20-year standard was financially benign when archives consisted solely of glass, and in our model its net cost for glass depended on late requests; by contrast, automatically extending a pre-digital 20-year retention rule to WSI imposes a substantial and recurring cost on laboratories adopting digital pathology.

The regulatory treatment of diagnostic digital images is now actively evolving. In October 2026, the United States Centers for Medicare & Medicaid Services and Centers for Disease Control and Prevention published a proposed rule under CLIA, not yet finalized, that would amend 42 CFR §493.1105(a)(7)(i)(B) to require clinical laboratories to retain both the histopathology glass slide and, if applicable, the digital slide image for 10 years, and §493.1101(e) to require that digital images be stored under conditions that ensure their preservation, noting that both media may be required to investigate complaints or quality issues.^8^ Similarly, the 2025 United Kingdom RCPath/IBMS guidance recommends retaining diagnostic WSI for 8 years, matching the retention period of replaceable glass slides, and explicitly states that digital images are not accepted as a substitute permitting early disposal of physical glass.^3^ By contrast, Australia’s National Pathology Accreditation Advisory Council (NPAAC) permits glass slides to be discarded after 4 years if the primary diagnosis was rendered on a WSI retained for 10 years,^9^ the CAP checklist position ties 10-year WSI retention to the unavailability of the original glass slides,^10^ and the 2014 CAP-ACP telepathology guideline defers WSI retention policy to regional requirements.^11^ If Canadian accrediting bodies or hospital policies adopt the dual-retention principle of the proposed CLIA rule and the RCPath/IBMS guidance by requiring WSI to be kept for the full duration of the glass slide requirement, a 20-year Canadian baseline would roughly double the image archive, and its storage cost, relative to a 10-year rule.

Our model indicates that Canadian laboratories and guideline bodies could avoid this cost escalation by decoupling digital image retention from physical glass and block retention, where clinical, accreditation, and legal requirements permit. If physical glass slides and paraffin blocks remain available for recutting or repeat scanning should a late clinical review, quality audit, or medicolegal inquiry arise, the economic case for maintaining petabyte-scale cloud WSI archives for two decades is weak; whether glass or a recut is an adequate substitute for the original digital record, including annotations, was not assessed. Empirical studies of digital pathology and radiology archives show that clinical image access decays steeply after the first year following diagnosis,^19–21^ although one radiology archive audit found that 5% of studies older than 10 years were opened over a 2-year period when they remained readily available.^22^ In our academic laboratory model, maintaining glass slides and blocks for 20 years while retaining WSI for 10 years reduced annual cost from CAD 569,308 to CAD 395,057, removing the second-decade cost increment at the base request share, while preserving 10 years of digital access for clinical care and 20 years of glass for late requests. Digital access from an archive tier is not immediate: restoring an image takes up to 15 hours at standard priority,^29^ so urgent requests would require faster restoration or a cached copy at additional cost. Placing post-diagnostic WSI in deep-archive cloud storage rather than hot storage kept digital retrieval costs (rehydration, 6-month hot caching, and egress) below 6% of the digital budget.

Selective WSI retention, archiving digital images for only a curated subset of cases, was the cheapest strategy in our base case only because we assumed that 10% of slides selected at sign-out would capture 90% of subsequent retrieval requests, a performance that no validated selection method has demonstrated. When evaluated against an illustrative imperfect capture curve (g = 3),^13,18,20^ selective WSI approximately broke even with glass-only retention, random selection increased net costs, and selection effort costing more than about CAD 0.28 per case would cancel the base-case saving. Selection by case and selection by slide also differ, particularly for large oncology cases. Laboratories contemplating selective digital archival should therefore audit their institutional requisition logs by specimen type, diagnosis, and case age before relying on selective retention to offset storage budgets.

Our results are consistent with prior operational and economic studies of pathology archives. Previous reviews have highlighted that personnel time represents a primary direct cost of physical slide archives,^12^ and institutional implementation series from Memorial Sloan Kettering Cancer Center and European centres have documented that digital WSI availability reduces physical slide room requests by 85% to 93%.^18,19,23^ While comprehensive business-case frameworks for digital pathology have identified long-term image storage as a major capital and operational expense,^14,15,34^ our study isolates the specific marginal cost of the retention rule itself and explains why the United Kingdom 8-year rule fails to reduce costs in a glass-only workflow: slides aged 8 and 9 years are requested frequently enough that the labour and histology costs of block recutting exceed the marginal savings in cabinet space. The off-site analysis points the same way. At published tariff rates, moving glass to a commercial vendor after the first year added roughly 43% to 62% to glass archive costs, because handling and retrieval, rather than storage space, dominate those costs; off-site storage is therefore justified by a shortage of hospital space rather than by savings.

Several limitations warrant consideration, most of which reflect the paucity of published data on pathology archive use. First, the second-decade request share is an illustrative assumption: we found no published study of how often glass aged 10 to 19 years is requested, and the recall data we cite describe cases recalled more than a year after diagnosis within a 4.5-year calendar window, not the second decade.^13,22^ Because the glass-only result changes direction near a 0.3% share, we report the full range and the break-even share. Second, recut and staining were costed from a US academic fee schedule used as a proxy; the glass-only result also reverses if this cost falls below about CAD 15 per slide at a 0.5% share, and a recut may not reproduce the original diagnostic material. Third, baseline retrieval demand (5 requests per 100 cases accessioned per year, 85% in the first year) was synthesized from tertiary-centre and digital access series rather than local logs, and the same slide count was used for accessioned and requested cases. Fourth, the selective WSI rule is hypothetical, the capture curves are illustrative, and the effort of selection was not costed. Fifth, labour times, the benefits load, productive time, cabinet prices, and cabinet life are assumptions, and floor space was valued at office rent as a proxy for hospital space. Sixth, our model excludes scanner acquisition, image management software, network upgrades, and active diagnostic caching, and therefore must not be interpreted as a total cost-of-ownership model for digital pathology. Seventh, cloud and vendor costs were calculated from public list prices on a single reference date; the off-site prices are a United States public-sector tariff, archive tiers differ in retrieval time and service terms, and enterprise discounts or future price changes would alter absolute digital costs, although the 2-fold volume ratio between a 20-year and a 10-year steady-state digital archive will persist. Eighth, storage of paraffin blocks was not modelled. Finally, this economic evaluation does not adjudicate the clinical, educational, or medicolegal adequacy of any specific retention duration. For these reasons the results should be read as scenario and threshold analyses that laboratories can update with local data, not as estimates of costs in any particular laboratory.

## Conclusion

Under the modelled assumptions, whether extending glass retention from 10 to 20 years adds or saves money depends on how often glass is requested in its second decade and on the cost of recutting discarded material; at the illustrative base values, avoided recuts offset the additional cabinet and space costs, and the result reverses below a second-decade request share of about 0.32%. Extending universal 40× WSI retention from 10 to 20 years increased archive costs by 35% to 37% in the base case, predominantly through storage charges. Separate retention periods for glass and images, and selective WSI retention, warrant further evaluation, with local measurement of retrieval patterns by specimen age and of selection performance, and with clinical, accreditation, and legal requirements considered separately.

## Supporting information

Supplementary File 1: Cost model workbook

Supplementary File 2: CHEERS 2022 checklist

## Acknowledgments

None.

## Author contributions

M.J.C. conceived the study, built the cost model, performed the analysis and wrote the manuscript. R.T.-L., D.B. and G.M.Y. contributed to the work and critically reviewed the manuscript. All authors approved the final version.

## Ethics statement

Not applicable. This study used published data and publicly available prices and involved no human participants, patient data or tissue; research ethics board review was not required.

## Competing interests

The authors declare no competing interests.

## Funding

The authors received no financial support for the research, authorship, and/or publication of this article.

## Data availability

All inputs and results of the cost model are provided in the supplementary files. No patient data were used.

## Use of artificial intelligence

Claude Opus 5.5 and Claude Fable 5.2 (Anthropic) were used to assist with model coding, reference checking, and drafting, editing and reviewing the manuscript and supplementary files. All authors reviewed the final versions and take full responsibility for the content. No patient data were provided to any tool.

## Notes

### Competing Interest Statement

The authors have declared no competing interest.

