## Supplementary File 2: CHEERS 2022 checklist for "A Cost Analysis of 10-Year Versus 20-Year Glass Slide Retention in the Era of Whole Slide Imaging"

*A Cost Analysis of 10-Year Versus 20-Year Glass Slide Retention in the Era of Whole Slide Imaging*

Topics and abbreviated item wording are carried over from the checklist of the previous version, which abbreviated Husereau D, et al. BMJ. 2022;376:e067975; they are not the verbatim CHEERS text. Locations refer to section headings, paragraph numbers within a section, tables and figures of the anonymized manuscript, to the title page and to the sheets of Supplementary File 1. Page numbers are not given because they depend on the journal layout.

| **Section** | **Topic** | **No.** | **Guidance (abbreviated)** | **Location where reported** |
| --- | --- | --- | --- | --- |
| Title | Title | 1 | Identify the study as an economic evaluation and specify the interventions being compared. | Title (identifies a cost analysis of 10 and 20 year retention rules for glass slides and whole slide images) |
| Abstract | Abstract | 2 | Provide a structured summary that highlights context, key methods, results, and alternative analyses. | Abstract; structured: Background, Methods, Results, Conclusions |
| Introduction | Background and objectives | 3 | Give the context for the study, the study question, and its practical relevance for decision making in policy or practice. | Introduction, paragraphs 1 to 4; study question in the last paragraph |
| Methods | Health economic analysis plan | 4 | Indicate whether a health economic analysis plan was developed and where available. | No health economic analysis plan was developed or registered; the model and its inputs are fully specified in Methods and in Supplementary File 1 (README, Inputs sheets) |
| Methods | Study population | 5 | Describe characteristics of the study population (such as age range, demographics, socioeconomic, or clinical characteristics). | Methods, Laboratory Tiers, Archival Scenarios, and Retention Policies; three laboratories of 100,000, 500,000 and 750,000 slides per year; no patient population |
| Methods | Setting and location | 6 | Provide relevant contextual information that may influence findings. | Introduction, paragraphs 1 to 2; Methods, Resource Valuation and Unit Costs; Canadian cloud regions; Ontario labour costs |
| Methods | Comparators | 7 | Describe the interventions or strategies being compared and why chosen. | Methods, Laboratory Tiers, Archival Scenarios, and Retention Policies; three scenarios, each under 10 and 20 year rules; United Kingdom 8 year rule and mixed rules; Table 2; off-site glass storage as a secondary analysis (Methods, Secondary Analysis: Off-Site Glass Storage; Table 3); Supplementary File 3 (Supplementary Table S7, Supplementary Table S8, Supplementary Table S10) |
| Methods | Perspective | 8 | State the perspective(s) adopted by the study and why chosen. | Methods, Study Design and Reporting Standards; laboratory perspective; economic costs including allocated staff time, annualized cabinet capital and the opportunity cost of floor space |
| Methods | Time horizon | 9 | State the time horizon for the study and why appropriate. | Methods, Outcome Measures and Sensitivity Analyses; primary steady state annual cost, independent of horizon; secondary net present cost over 30 years from an empty archive |
| Methods | Discount rate | 10 | Report the discount rate(s) and reason chosen. | Methods, Outcome Measures and Sensitivity Analyses; 1.5%; 0% and 3% in sensitivity analysis; Supplementary File 3 (Supplementary Table S6, Supplementary Table S9) |
| Methods | Selection of outcomes | 11 | Describe what outcomes were used as the measure(s) of benefit(s) and harm(s). | Not applicable: cost analysis; no health outcomes. Cost outcomes and request counts (glass pulls, recuts, digital restores) are defined in Methods, Outcome Measures and Sensitivity Analyses |
| Methods | Measurement of outcomes | 12 | Describe how outcomes used to capture benefit(s) and harm(s) were measured. | Not applicable: no health outcomes measured; request counts derived from per case request rates in Methods, Clinical Retrieval Demand and Block Recutting |
| Methods | Valuation of outcomes | 13 | Describe the population and methods used to measure and value outcomes. | Not applicable: outcomes were not valued (cost analysis) |
| Methods | Measurement and valuation of resources and costs | 14 | Describe how costs were valued. | Methods, Resource Valuation and Unit Costs; Supplementary File 3 (Supplementary Table S1, Supplementary Table S2); Supplementary File 1 (Inputs, Derived sheets) |
| Methods | Currency, price date, and conversion | 15 | Report the dates of the estimated resource quantities and unit costs, plus the currency and year of conversion. | Methods, Study Design and Reporting Standards; 2026 CAD; 1.4257 CAD per USD, Bank of Canada, 7 October 2026; Supplementary File 3 (Supplementary Table S1); Discussion, paragraph 7 (limitations): list prices on one date; 2025 US vendor prices converted at the 2026 exchange rate without inflation adjustment |
| Methods | Rationale and description of model | 16 | If modelling is used, describe in detail and why used. Report if the model is publicly available and where it can be accessed. | Methods, Study Design and Reporting Standards; Supplementary File 1 (README, Calc_SS, Calc_NPC sheets); Title page, Statements and Declarations (Data availability) |
| Methods | Analytics and assumptions | 17 | Describe any methods for analysing or statistically transforming data, any extrapolation methods, and approaches for validating any model used. | Methods, Outcome Measures and Sensitivity Analyses; Methods, Clinical Retrieval Demand and Block Recutting; steady state, discounting, request age profile, recut share and selective WSI assumptions; assumptions to confirm in Supplementary File 3 (Supplementary Table S3); validation by cross checks against earlier model versions and an independent comparator model in Supplementary File 1 (CrossCheck, Comparator_check sheets) Every input is classified as a published value, a published value used as a proxy, or an assumption (Supplementary Table S1); the second-decade request share is an illustrative assumption reported with break-even thresholds. The workbook was recalculated from its inputs and its results reproduced by independent code (Supplementary File 1, README). |
| Methods | Characterising heterogeneity | 18 | Describe any methods used for estimating how the results of the study vary for subgroups. | three laboratory sizes in Methods, Laboratory Tiers, Archival Scenarios, and Retention Policies; Table 1; Figure 1; Figure 2; no patient subgroups |
| Methods | Characterising distributional effects | 19 | Describe how impacts are distributed across different individuals or adjustments made to reflect priority populations. | Not applicable: laboratory cost analysis; no effects on individuals or priority populations were modelled |
| Methods | Characterising uncertainty | 20 | Describe methods to characterise any sources of uncertainty in the analysis. | Methods, Outcome Measures and Sensitivity Analyses; one way deterministic; probabilistic analysis not performed; Results, Sensitivity Analyses; Supplementary File 3 (Supplementary Table S5, Supplementary Table S6, Supplementary Figure S2); Supplementary File 1 (Sensitivity sheet) Two-way break-even thresholds for request share against recut cost and for selective coverage against selection effort (Supplementary Table S11, Supplementary Figure S3); cloud tier, 20x scanning and off-site storage reported as scenarios. |
| Methods | Approach to engagement with patients and others affected by the study | 21 | Describe any approaches to engage patients or service recipients, the general public, communities, or stakeholders in the design of the study. | No patients, members of the public or stakeholders were engaged in the design (cost analysis of published data and public prices) |
| Results | Study parameters | 22 | Report all analytic inputs (such as values, ranges, references) including uncertainty or distributional assumptions. | Methods, Clinical Retrieval Demand and Block Recutting; Methods, Resource Valuation and Unit Costs; Supplementary File 3 (Supplementary Table S1, Supplementary Table S2, Supplementary Table S3); Supplementary File 1 (Inputs sheet) |
| Results | Summary of main results | 23 | Report the mean values for the main categories of costs and outcomes of interest and summarise them in the most appropriate overall measure. | Results, Scenarios 1 to 3; Results, Decoupled Media Retention Policies and Jurisdiction Comparisons; Results, Off-Site Glass Storage (Secondary Analysis); Table 3; Figure 3; Table 1; Table 2; Figure 1; Figure 2; Supplementary File 3 (Supplementary Table S4, Supplementary Figure S1); Supplementary File 1 (Results_3x3 sheet) |
| Results | Effect of uncertainty | 24 | Describe how uncertainty about analytic judgments, inputs, or projections affect findings. Report the effect of choice of discount rate and time horizon, if applicable. | Results, Sensitivity Analyses; 30 year net present cost; Supplementary File 3 (Supplementary Table S5, Supplementary Table S6, Supplementary Table S9, Supplementary Figure S2); Supplementary File 1 (Sensitivity sheet) Break-even thresholds: Results, Sensitivity Analyses; Supplementary Table S11; Supplementary Figure S3. |
| Results | Effect of engagement with patients and others affected by the study | 25 | Report on any difference patient/service recipient, general public, community, or stakeholder involvement made to the approach or findings of the study. | None: no engagement was undertaken (see item 21) |
| Discussion | Study findings, limitations, generalisability, and current knowledge | 26 | Report key findings, limitations, ethical or equity considerations not captured, and how these could affect patients, policy, or practice. | Discussion, paragraphs 1 to 7; limitations in paragraph 7; Conclusion |
| Other relevant information | Source of funding | 27 | Describe how the study was funded and any role of the funder in the identification, design, conduct, and reporting of the analysis. | Title page, Statements and Declarations (Funding) |
| Other relevant information | Conflicts of interest | 28 | Report authors’ conflicts of interest according to journal or International Committee of Medical Journal Editors requirements. | Title page, Statements and Declarations (Declaration of conflicting interest) |
