## Supplementary File 3: Supplementary tables and figures for "A Cost Analysis of 10-Year Versus 20-Year Glass Slide Retention in the Era of Whole Slide Imaging"

**Supplementary File 3. Detailed inputs, assumptions, results and sensitivity analyses**

Cost of retaining glass slides and whole slide images under 10 and 20 year rules for three laboratory sizes. Every number in this file is read by script from the recalculated cost model (Supplementary File 1; sheet and cell list in the build log). Currency: Canadian dollars (CAD) 2026; USD prices converted at 1.4257 CAD per USD inside the model. Values are steady state annual costs with a full archive unless stated. Version 7: base second-decade request share 0.5% (an illustrative assumption; 0.01% in version 5, kept as a sensitivity value); every input carries an evidence class (Table S1); Table S10 gives commercial off-site glass storage and Table S11 the break-even thresholds; Supplementary Figure S3 shows the threshold analyses. Laboratories: small community 100,000, academic 500,000 and large academic 750,000 slides per year. Scenarios: S1 glass only; S2 glass plus whole slide images (WSI) of every slide; S3 glass plus selective WSI (10% of slides assumed to cover 90% of digitally servable requests). Negative numbers are written with a hyphen minus sign. Terms: the source evidence dossier ("dossier") is the set of research briefs, literature review and three prior cost models assembled for this study (prior models S1 to S3: digital archive storage, glass slide storage and selective digital coverage); "study specification" denotes scenario definitions set by the study authors, which are author assumptions. Revised November 25, 2026: the cabinet price input was updated from USD 169 to the USD 199 list price shown on the cited product page, and every result in this file was recalculated.

Table S1. Model inputs, values, ranges, sources and status

Table S2. Archive tier price table (Canadian regions)

Table S3. Inputs that are author or analyst assumptions to be confirmed

Table S4. Full results by laboratory, scenario, retention rule and cost component

Table S5. Selective WSI: selection quality and break even coverage

Table S6. One way sensitivity analysis, all laboratories

Table S7. Annual cost under the 8, 10 and 20 year jurisdiction rules

Table S8. Mixed retention rules (different periods for glass and images)

Table S9. Secondary outcome: net present cost over 30 years

Table S10. Secondary analysis: commercial off-site storage of glass slides

Table S11. Break-even thresholds for the glass-only retention decision and for selective WSI

Supplementary Figure S1. Annual cost under the 8, 10 and 20 year jurisdiction rules

Supplementary Figure S2. Selective WSI: cost relative to glass only under alternative selection rules

Supplementary Figure S3. Break-even thresholds

**Table S1. Model inputs, values, ranges, sources and status (Supplementary File 1, sheet Inputs)**

| **Name** | **Description** | **Value** | **Unit** | **Sensitivity range** | **Public source (v7)** | **Evidence class (v7)** | **Confirm** |
| --- | --- | --- | --- | --- | --- | --- | --- |
| **A. Laboratories, currency, discounting, horizon** | | | | | | | |
| VOL_S | Small community laboratory: glass slides produced and archived per year | 100,000 | slides/yr |  | Authors; illustrative laboratory sizes, not survey data | Study specification (illustrative) | N |
| VOL_A | Academic laboratory | 500,000 | slides/yr |  | Authors; illustrative laboratory sizes, not survey data | Study specification (illustrative) | N |
| VOL_L | Large academic laboratory | 750,000 | slides/yr |  | Authors; illustrative laboratory sizes, not survey data | Study specification (illustrative) | N |
| SPC | Slides per case = slides per glass request (one case-level request); cases/yr = slides/yr / SPC | 3 | slides/case | 1 to 10 | Authors; the same value is used for slides per accessioned case and slides per requested case | Assumption | Y |
| FX | Bank of Canada FXUSDCAD 2026-10-07; one rate for every USD price; USD = CAD / FX | 1.4257 | CAD per USD |  | Bank of Canada FXUSDCAD, 7 October 2026 (manuscript ref 17) | Published value | N |
| DISC | Discount rate, CDA-AMC reference case; NPC = sum C_t / (1+d)^(t-1); also annualizes cabinets (CRF) | 0.015 | per yr (real) | 0 to 0.03 | CADTH 2017 guidelines, 1.5% reference case (ref 32) | Published convention | N |
| DISC_LO | Low discount rate (sensitivity) | 0 | per yr |  | CADTH 2017 guidelines, sensitivity (ref 32) | Published convention | N |
| DISC_HI | High discount rate (sensitivity) | 0.03 | per yr |  | CADTH 2017 guidelines, sensitivity (ref 32) | Published convention | N |
| H_NPC | Horizon of the secondary metric (NPC from an empty archive): 30 y lets a 20-y rule run 10 years at steady state | 30 | years |  | Authors | Model convention | N |
| **B. Retention periods and demand age bands** | | | | | | | |
| RG_SHORT | Short retention rule, 10 y (US CLIA/CAP; in Canada, the Ontario regulatory minimum and OAML adult slides, which conflict with the CAP-ACP 20 y guidance) | 10 | years |  | CLIA 42 CFR 493.1105 (ref 1); OAML CLP020-001 (ref 7) | Published retention rule | N |
| RG_LONG | Long retention rule (CAP-ACP 2005; BC PLMS 2024) | 20 | years |  | CAP-ACP 2005 (ref 4); BC PLMS 2024 (ref 5) | Published retention guidance | N |
| RG_UK | UK RCPath/IBMS G031 2025: minimum 8 y if the slide is replaceable from the block (else 30 y) | 8 | years |  | RCPath/IBMS G031 2025 (ref 3); UK block and irreplaceable-slide rules not modelled | Published retention guidance; modelled as an 8-year scenario only | N |
| AGE_SPLIT | Requests for cases aged 0 to 9 follow P0; ages 10 to 19 carry the recut-share tail | 10 | years |  | Authors | Model structure | N |
| AGE_MAX | No request modelled for cases aged 20 y or more (block retention 20 y, OAML adult blocks and CAP-ACP) | 20 | years |  | Authors; blocks assumed kept 20 years in every policy | Model structure | N |
| **C. Glass: labour, loss and recut (S2 Assumptions)** | | | | | | | |
| WAGE | Medical laboratory assistant median wage, an Ontario economic region, Job Bank NOC 33101 | 28.14 | CAD/h (2024) | 22.5 to 34.57 | Job Bank NOC 33101 medical laboratory assistant, London region median, updated 19 Nov 2025 (ref 24) | Published value | N |
| CPI_NOW | Canada CPI all-items, Aug 2026 | 169.8 | index |  | Statistics Canada CPI all-items (Table 18-10-0004-01), August 2026 | Published value | N |
| CPI_WAGE | Canada CPI 2024 annual average (wage reference year) | 160.85 | index |  | Statistics Canada CPI all-items, 2024 annual average | Published value | N |
| EBP | Benefits load (Treasury Board proxy) | 0.27 | fraction | 0.2 to 0.35 | 27% benefits load; federal public-service proxy, not hospital-specific | Assumption (proxy) | N |
| PROD | Productive share of paid hours | 0.85 | fraction | 0.75 to 0.95 | Authors | Assumption | Y |
| FILE_MIN | On-site filing per slide (one-time, rule-invariant) | 0.25 | min/slide | 0.1 to 0.5 | Authors; no time-motion data | Assumption | Y |
| DESTR_MIN | On-site culling per slide at end of retention (rule-invariant at steady state) | 0.05 | min/slide | 0.02 to 0.2 | Authors; no time-motion data | Assumption | Y |
| PULL_MIN | Locate, pull and check out a case | 8 | min/request |  | Authors; no time-motion data | Assumption | Y |
| REFILE_MIN | Re-file returned slides | 4 | min/request |  | Authors; no time-motion data | Assumption | Y |
| TRACK_MIN | Requisition logging and tracking | 3 | min/request |  | Authors; no time-motion data | Assumption | Y |
| ONSITE_MIN_LO | On-site labour per request, low (Eccher 2023 about 10 min per case) | 10 | min/request |  | Eccher 2023, about 10 minutes per case (ref 12) | Assumption (literature-informed) | N |
| ONSITE_MIN_HI | On-site labour per request, high (Hanna 2019 calibration) | 20 | min/request |  | Authors | Assumption | N |
| LOSS_P | Probability a loaned slide is lost (Giannini 2011) | 0.0002 | per slide loaned |  | Published slide-loss estimate (Giannini 2011; not cited in the manuscript) | Assumption (literature-informed) | N |
| REPL_USD | Recut plus H&E per slide (UCLA TPCL 7.30 + 8.80) | 16.1 | USD/slide |  | UCLA TPCL internal rates, recut USD 7.30 + H&E USD 8.80, effective April 2026 (ref 25); US academic recharge, not a Canadian marginal cost | Published value used as proxy | N |
| BLOCK_PULL | 1 = a recut request also pays one block pull, costed like a glass pull (dossier brief 5.6: 'the cost of pulling a glass slide and pulling a block are similar'); 0 = comparator convention (recut replaces the pull) | 1 | 0/1 |  | Authors; a block pull is costed like a glass pull | Assumption | Y |
| RECUT_ON | 1 = requests for discarded glass (ages RG to AGE_MAX-1) are met by recut from the retained block and costed (study specification); 0 = not costed (prior models S2/S3 base convention) | 1 | 0/1 |  | Authors | Model structure | N |
| **D. Glass: on-site space and cabinets (S2 Assumptions)** | | | | | | | |
| SQFT_PER_M2 | Unit conversion | 10.7639 | sq ft/m2 |  |  | Exact conversion | N |
| RENT_OFF_NET | Office net asking rent, Q2 2026 (mid-sized Ontario city; proxy for opportunity cost of hospital space) | 13.77 | CAD/sq ft/yr |  | Cushman & Wakefield office MarketBeat Q2 2026 (ref 27); office asking rent as a proxy for hospital space | Published value used as proxy | N |
| RENT_OFF_ADD | Office additional rent, Q2 2026 | 12.41 | CAD/sq ft/yr |  | Cushman & Wakefield office MarketBeat Q2 2026 (ref 27) | Published value used as proxy | N |
| RENT_IND | Industrial gross rent (low space case) | 14.78 | CAD/sq ft/yr |  | Industrial rent, MarketBeat Q2 2026 (not cited in the manuscript) | Published value used as proxy (sensitivity) | N |
| HOSP_PSF_LO | New hospital hard cost, GTA, low | 1,030 | CAD/sq ft |  | Published construction-cost range, Greater Toronto Area (not cited in the manuscript) | Published value used as proxy (sensitivity) | N |
| HOSP_PSF_HI | New hospital hard cost, GTA, high | 1,620 | CAD/sq ft |  | Published construction-cost range, Greater Toronto Area (not cited in the manuscript) | Published value used as proxy (sensitivity) | N |
| BLDG_LIFE | Building life for annualizing construction | 40 | years |  | Authors | Assumption | Y |
| SPACE_SEL | Space cost case: 1 industrial rent, 2 office gross rent (base), 3 new hospital build | 2 | 1/2/3 | 1 to 3 | Authors | Analyst choice | N |
| CAB_CAP | SS-200 cabinet capacity | 5,000 | slides |  | Phoenix SS-200 product page (ref 26) | Published specification | N |
| CAB_W | Cabinet width | 0.425 | m |  | Phoenix SS-200 (ref 26) | Published specification | N |
| CAB_D | Cabinet depth | 0.476 | m |  | Phoenix SS-200 (ref 26) | Published specification | N |
| STACK_N | Cabinets per stack | 12 | cabinets | 8 to 15 | Authors; stack height | Assumption | N |
| FILL | Usable fill of cabinet capacity | 0.85 | fraction | 0.75 to 0.95 | Authors | Assumption | Y |
| AISLE_W | Aisle width | 1.1 | m | 0.9 to 1.5 | Authors | Assumption | Y |
| CIRC | Circulation allowance | 0.15 | fraction | 0.1 to 0.25 | Authors | Assumption | Y |
| CAB_PRICE_USD | SS-200 cabinet list price | 199 | USD each |  | Phoenix SS-200 product page (ref 26); list price of USD 199 shown on November 25, 2026 (USD 169 in version 7) | Published list price | N |
| BASE_PRICE_USD | SB-200 base list price | 69 | USD each |  | Phoenix SB-200 base (ref 26); price not shown on the product page | Vendor price not confirmed on the cited page (treat as assumption) | N |
| CAB_DISC | Cabinet volume discount | 0.1 | fraction |  | Authors; assumed volume discount | Assumption | N |
| BASE_DISC | Base volume discount | 0.1 | fraction |  | Authors; assumed volume discount | Assumption | N |
| LANDED | Landed-cost uplift (freight, brokerage, tax) | 0.25 | fraction | 0.1 to 0.4 | Authors | Assumption | Y |
| CAB_LIFE | Cabinet life for annualizing | 20 | years | 10 to 30 | Authors | Assumption | Y |
| STACK_ROUND | 1 = cabinets and floor space bought in whole stacks (stepwise, study specification); 0 = continuous per slide-year (dossier convention) | 1 | 0/1 | 0 to 1 | Authors | Model convention | N |
| **E. Glass: off-site vendor (Iron Mountain US public-sector tariff, pathology lines; sensitivity only)** | | | | | | | |
| PATH_STOR | Temperature-controlled storage | 2.55 | USD/cu ft/month |  | Iron Mountain Attachment D, 2025 (ref 33), row 594; US public-sector price, not a Canadian quotation | Published US tariff used as proxy | N |
| PATH_RECV | Receiving and entry | 6.57 | USD/case |  | Iron Mountain Attachment D, 2025 (ref 33), row 595; US public-sector price, not a Canadian quotation | Published US tariff used as proxy | N |
| PATH_LIST | Individual listing | 0.7 | USD/case |  | Iron Mountain Attachment D, 2025 (ref 33), row 603; US public-sector price, not a Canadian quotation | Published US tariff used as proxy | N |
| PATH_RET | Regular retrieval | 3.51 | USD/case |  | Iron Mountain Attachment D, 2025 (ref 33), row 596; US public-sector price, not a Canadian quotation | Published US tariff used as proxy | N |
| PATH_REFILE | Regular refile | 3.51 | USD/case |  | Iron Mountain Attachment D, 2025 (ref 33), row 598; US public-sector price, not a Canadian quotation | Published US tariff used as proxy | N |
| PATH_DEL | Next-day delivery | 42.87 | USD/visit |  | Iron Mountain Attachment D, 2025 (ref 33), row 606; US public-sector price, not a Canadian quotation | Published US tariff used as proxy | N |
| PATH_PICKUP | Regular pick-up | 66.6 | USD/visit |  | Iron Mountain Attachment D, 2025 (ref 33), row 607; US public-sector price, not a Canadian quotation | Published US tariff used as proxy | N |
| PATH_HANDLING | Handling, charged once at intake | 4.11 | USD/cu ft |  | Iron Mountain Attachment D, 2025 (ref 33), row 613; US public-sector price, not a Canadian quotation | Published US tariff used as proxy | N |
| PATH_ADMIN | Administrative fee | 30.26 | USD/month |  | Iron Mountain Attachment D, 2025 (ref 33), row 614; US public-sector price, not a Canadian quotation | Published US tariff used as proxy | N |
| IM_MIN_MONTH | Monthly minimum storage billing | 341.33 | USD/month |  | Iron Mountain Attachment D, 2025 (ref 33), row 605; US public-sector price, not a Canadian quotation | Published US tariff used as proxy | N |
| IM_DESTR_CF | Destruction per carton volume | 3.78 | USD/cu ft |  | Iron Mountain Attachment D, 2025 (ref 33), row 48 (records section); US public-sector price, not a Canadian quotation | Published US tariff used as proxy | N |
| IM_RET_CARTON | Carton retrieval added to destruction | 4.11 | USD/cu ft |  | Iron Mountain Attachment D, 2025 (ref 33), row 15 (records section); US public-sector price, not a Canadian quotation | Published US tariff used as proxy | N |
| SLIDES_PER_BOX | Slides per cardboard slide box | 2,000 | slides |  | Slide storage box and carton catalogues | Vendor catalogue value (not cited in the manuscript) | N |
| CARTON_CF | Billed volume per carton | 1.2 | cu ft |  | Iron Mountain Attachment D, 2025 (ref 33), row records carton volume; US public-sector price, not a Canadian quotation | Published US tariff used as proxy | N |
| TEK_PRICE | Slide file case (10 boxes) | 277 | USD/case |  | Slide storage box and carton catalogues | Vendor catalogue value (not cited in the manuscript) | N |
| TEK_SLIDES | Slides per slide file case | 20,000 | slides |  | Slide storage box and carton catalogues | Vendor catalogue value (not cited in the manuscript) | N |
| CARTON_PRICE | Standard carton | 5.67 | USD each |  | Slide storage box and carton catalogues | Vendor catalogue value (not cited in the manuscript) | N |
| INTAKE_TRIPS | Transfer pick-ups per year | 12 | trips/yr |  | Authors | Assumption | Y |
| BOX_PER_CARTON | Slide boxes per carton | 2 | boxes |  | Authors | Assumption | Y |
| OFF_FILL | Usable fill of off-site boxes | 0.85 | fraction |  | Authors | Assumption | Y |
| EV_PER_TRIP | Requests sharing one delivery and pick-up trip | 20 | requests/trip | 5 to 50 | Authors | Assumption | Y |
| OFF_INHOUSE_MIN | In-house labour per off-site request | 6 | min/request | 4 to 10 | Authors | Assumption | Y |
| GLASS_SITE | Glass archive used in the base: 1 = on site (base), 2 = off-site vendor (sensitivity) | 1 | 1/2 | 1 to 2 | Authors | Analyst choice | N |
| **F. Request rate (identical in every scenario; S3 coverage model, literature.md section 4.2)** | | | | | | | |
| P0_RATE | Glass requests per year per 100 cases accessioned per year, cases aged 0 to 9 y, no digital archive (LabPON about 2.7, MSKCC 2014 about 12) | 5 | requests per 100 cases/yr | 2.5 to 12 | Synthesized from MSK (about 12 per 100 cases, Hanna 2019, ref 18) and LabPON (about 2.7, Baidoshvili 2018, ref 19); no Canadian data | Assumption (literature-informed) | Y |
| P0_LO | Low request rate | 2.5 | per 100 cases/yr |  | Refs 18 and 19 | Sensitivity value | N |
| P0_HI | High request rate (MSKCC 2014) | 12 | per 100 cases/yr |  | Hanna 2019 (ref 18) | Sensitivity value | N |
| A1_SHARE | Share of age 0 to 9 demand falling in the first 12 months; rest spread evenly over ages 1 to 9 | 0.85 | share | 0.75 to 0.95 | Shape informed by first-access curves after scanning (Schüffler 2021, ref 20) and radiology (refs 21, 22); no measured age distribution of glass requests | Assumption (literature-informed) | Y |
| P100_RATE | Residual glass requests when the case has a retained WSI (floor; about 90% fall, MSK 93%) | 0.5 | per 100 cases/yr | 0.2 to 1.7 | Hanna 2019: 93% fewer glass requests once images were available (ref 18) | Assumption (literature-informed) | Y |
| P100_LO | Low residual glass need | 0.2 | per 100 cases/yr |  | Authors | Sensitivity value | N |
| P100_HI | High residual glass need (Ardon 2021 implies up to about 34%) | 1.7 | per 100 cases/yr |  | Ardon 2023 (ref 23) | Sensitivity value | N |
| **G. Recut share for glass discarded at 10 years (study specification, scenario 1)** | | | | | | | |
| RECUT_SHARE | BASE recut share: share of one year's slides (equivalently cases, SPC slides per request) that are requested once while aged 10 to 19 y. Steady-state recut requests per year = RECUT_SHARE x cases/yr. Per case-year request rate at ages 10 to 19 = RECUT_SHARE / 10. v6 base 0.5% (0.05% per case-yr), anchored to Ali 2023; v5 used 0.01%. No pathology source measures requests for 10 to 20 y glass | 0.005 | share of a year's slides |  | No published data on requests for 10 to 20-year-old glass. 0.5% is illustrative, informed by Ali 2023 (ref 13: 0.24% of cases recalled more than 12 months after diagnosis), which is not an age-specific rate; the 0.29% break-even is reported | Illustrative assumption (no data) | Y |
| **H. Selective WSI (scenario 3)** | | | | | | | |
| SEL_SHARE | Share of each year's slides whose WSI is retained (selected at sign-out) | 0.1 | share of slides | 0.05 to 0.2 | No validated selection rule; 10% of slides covering 90% of requests is hypothetical | Study specification (hypothetical) | Y |
| SEL_CAPTURE | Share of digitally servable requests that fall on the retained subset (coverage). Requests on covered cases still need glass at the floor share PHI, as in S2 | 0.9 | share of requests | 0.8 to 0.95 | No validated selection rule; capture refers to digitally servable requests | Study specification (hypothetical) | Y |
| SEL_SHARE_LO | Coverage sensitivity pair 1: share | 0.05 | share |  |  | Sensitivity value | N |
| SEL_CAP_LO | Coverage sensitivity pair 1: coverage | 0.8 | share |  |  | Sensitivity value | N |
| SEL_SHARE_HI | Coverage sensitivity pair 3: share | 0.2 | share |  |  | Sensitivity value | N |
| SEL_CAP_HI | Coverage sensitivity pair 3: coverage | 0.95 | share |  |  | Sensitivity value | N |
| G_BASE | Literature-calibrated capture curve C(r) = 1-(1-r)^g, imperfect prediction (base of prior model S3) | 3 | exponent |  | Illustrative capture curve; the derivation is not reproducible from published data | Illustrative assumption | N |
| G_OPT | Literature-calibrated capture curve, better prediction (MSK about 6.9, Coventry M-code about 8.1) | 7.5 | exponent |  | Illustrative capture curve; the derivation is not reproducible from published data | Illustrative assumption | N |
| **I. Digital archive (S1 digital model; all prices are Canadian regions, PHIPA/PIPEDA-compliant per dossier; list prices accessed 2026-10-08)** | | | | | | | |
| GB_40X | WSI size at 40x | 1.5 | GB/slide |  | Plausible 40x file size; published series report a range (refs 20, 23) | Assumption (literature-informed) | Y |
| GB_20X | WSI size at 20x (sensitivity) | 0.5 | GB/slide |  | Authors | Assumption (scenario) | N |
| OBJ | Storage objects per WSI | 1 | objects/slide |  | Authors | Model convention | N |
| TIER_SEL | Base archive tier: 1 Azure Archive LRS 3-y reserved (Canada Central); 2 AWS S3 Glacier Deep Archive list (ca-central-1); 3 Azure Archive LRS pay-as-you-go; 4 Google Cloud Archive (Montreal/Toronto) | 1 | 1 to 4 | 1 to 4 | Lowest-priced evaluated Canadian-region archive tier | Analyst choice | N |
| HOLD_M | Hot storage of each restored case before deletion (study specification: 6 months, not 90 days) | 6 | months | 3 to 12 | Authors | Study specification (assumption) | N |
| HOLD_M_LO | Hot hold sensitivity, low | 3 | months |  |  | Sensitivity value | N |
| HOLD_M_HI | Hot hold sensitivity, high | 12 | months |  |  | Sensitivity value | N |
| EGRESS_SEL | Egress of each restored case once to the hospital: 1 internet egress tiers (base, conservative); 2 dedicated link (DTO only, port and circuit fees excluded); 0 none (viewer in the same cloud region) | 1 | 0/1/2 | 0 to 2 | Authors | Analyst choice | Y |
| RESTORE_SEL | Restore volume: 1 = request-driven (each digitally served request restores its case, SPC x GB); 2 = prior model S1 convention (RECALL_D of the archive per year) | 1 | 1/2 | 1 to 2 | Authors | Analyst choice | Y |
| RECALL_D | Prior model S1 recall convention (sensitivity and cross-check only) | 0.05 | share of archive/yr |  | Authors | Earlier-model convention (sensitivity only) | N |
| HOLD_D_PKG | Prior model S1 hot hold (cross-check only; v3 uses HOLD_M) | 90 | days |  | Authors | Earlier-model convention (not used) | N |
| AZR_1PB_GB | Azure reservation unit, 1 PB | 1,048,576 | GB |  | Azure reservation unit, 1 PiB = 1,048,576 GiB (binary units) | Unit definition | N |
| AZR_100TB_GB | Azure reservation unit, 100 TB | 102,400 | GB |  | Azure reservation unit, 100 TiB = 102,400 GiB (binary units) | Unit definition | N |
| AZR_1PB_P | Archive LRS 1 PB 3-year reservation, Canada Central | 52,999 | USD per 3-y term |  | Azure Retail Prices API, Archive LRS 3-year reservation, Canada Central, accessed 8 Oct 2026 (ref 28) | Published list price | N |
| AZR_100TB_P | Archive LRS 100 TB 3-year reservation, Canada Central (minimum commitment unit) | 5,441 | USD per 3-y term |  | Azure Retail Prices API, as above (ref 28) | Published list price | N |
| AZR_TERM | Reservation term | 3 | years |  | Azure reservation term (ref 28) | Published list price | N |
| **J. Jurisdiction retention periods for adult surgical glass slides (years; Jurisdictions sheet)** | | | | | | | |
| RET_US | United States: CLIA 42 CFR 493.1105 (at least 10 y from examination); CAP (US) 2010 10 y | 10 | years |  | 42 CFR 493.1105 (ref 1); CAP 2010 (ref 2) | Published retention rule | N |
| RET_UK | United Kingdom: RCPath/IBMS G031 6th ed. 2025, minimum 8 y if replaceable from the block, else 30 y | 8 | years |  | RCPath/IBMS G031 2025 (ref 3) | Published retention guidance | N |
| RET_ON | Ontario: OAML CLP020-001 (2006) citing OLA, adult slides 10 y (blocks 20 y); Reg. 965 s. 20 minimum 10 y | 10 | years |  | Reg. 965 s. 20 (ref 6; 2011 consolidation); OAML 2006 (ref 7) | Published retention rule (current text not re-verified) | N |
| RET_CAPACP | Canada: CAP-ACP 'Retention and use of human biologic material' (approved Nov 2005), 20 y | 20 | years |  | CAP-ACP 2005 (ref 4) | Published retention guidance | N |
| RET_BC | British Columbia: PLMS guideline v2.2 (July 2024), 20 y | 20 | years |  | BC PLMS v2.2, 2024 (ref 5) | Published retention guidance | N |
| RET_AUS | Australia: NPAAC 9th ed. 2022, slides 10 y (4 y if the diagnosis is made on a digital image kept 10 y) | 10 | years |  | NPAAC 9th ed. 2022 (ref 9) | Published retention requirement | N |

Evidence class (v7): Published value = taken directly from the cited public source; Published value used as a proxy = a real published value applied to a setting in which it was not measured (for example a US tariff or US academic fee schedule applied to Canada, or office rent applied to hospital space); Assumption = set by the authors without a published source (literature-informed where noted); Illustrative assumption = a value chosen to illustrate the model where no data exist, reported with break-even thresholds; Study specification = scenario definitions set by the authors; Sensitivity value = alternative value used only in sensitivity analysis. Earlier internal model files that were cited as sources in previous versions are not supplied and are no longer used as provenance.

**Table S2. Archive tier price table (USD list prices, Canadian regions; Supplementary File 1, sheet Inputs, section K)**

| **Name** | **Description** | **Unit** | **1 Azure Archive LRS 3-y reserved, Canada Central** | **2 AWS S3 Glacier Deep Archive, ca-central-1** | **3 Azure Archive LRS pay-as-you-go, Canada Central** | **4 Google Cloud Archive, Montreal/Toronto** |
| --- | --- | --- | --- | --- | --- | --- |
| T_STOR | Archive storage list price (tier 1 is priced by reservation blocks, see AZR_*; value shown is the effective 1 PB block price, information only) | USD/GB-month | 0.001403994 | 0.0018 | 0.0018 | 0.0025 |
| T_RES | 1 = priced by Azure reservation blocks | 0/1 | 1 | 0 | 0 | 0 |
| T_PUT | Ingest write operations | USD/1,000 | 0.011 | 0.06 | 0.011 | 0.05 |
| T_RETR | Archive retrieval / rehydration (Azure standard up to 15 h; AWS bulk within 48 h) | USD/GB | 0.022 | 0.005 | 0.022 | 0.05 |
| T_REQ | Archive read / restore requests | USD/1,000 | 0.55 | 0.0275 | 0.55 | 0.05 |
| T_HOTPUT | Write of the restored copy to the hot tier | USD/1,000 | 0.0055 | 0.0055 | 0.0055 | 0.005 |
| T_HL1 | Hot tier: tier-1 limit (Azure Hot LRS row 7; S3 Standard row 2; GCS Standard flat row 14) | GB | 51,200 | 51,200 | 51,200 | 1,000,000,000,000,000 |
| T_HR1 | Hot tier: tier-1 price | USD/GB-month | 0.02 | 0.025 | 0.02 | 0.023 |
| T_HL2 | Hot tier: tier-2 limit | GB | 512,000 | 512,000 | 512,000 | 1,000,000,000,000,000 |
| T_HR2 | Hot tier: tier-2 price | USD/GB-month | 0.0192 | 0.024 | 0.0192 | 0.023 |
| T_HR3 | Hot tier: tier-3 price | USD/GB-month | 0.0184 | 0.023 | 0.0184 | 0.023 |
| T_E1L | Internet egress: tier-1 limit | GB/month | 10,335 | 10,240 | 10,335 | 10,240 |
| T_E1R | Internet egress: tier-1 price | USD/GB | 0.087 | 0.09 | 0.087 | 0.12 |
| T_E2L | Internet egress: tier-2 limit | GB/month | 51,295 | 51,200 | 51,295 | 153,600 |
| T_E2R | Internet egress: tier-2 price | USD/GB | 0.083 | 0.085 | 0.083 | 0.11 |
| T_E3L | Internet egress: tier-3 limit | GB/month | 153,695 | 153,600 | 153,695 | 153,600 |
| T_E3R | Internet egress: tier-3 price | USD/GB | 0.07 | 0.07 | 0.07 | 0.11 |
| T_E4R | Internet egress: price above tier 3 | USD/GB | 0.05 | 0.05 | 0.05 | 0.08 |
| T_DTO | Dedicated-link data transfer out (DTO only) | USD/GB | 0.025 | 0.02 | 0.025 | 0.02 |

Tier 1 (base case) storage price is the effective monthly price of a 3 year Azure reservation derived in the model from the reservation inputs in Table S1. Prices were taken from the source evidence dossier (list prices accessed 2026-10-08) and were not re-fetched. Very large limit values denote an unlimited tier in the source price table.

**Table S3. Inputs that are author or analyst assumptions to be confirmed (Confirm = Y in Table S1)**

| **Name** | **Description** | **Value** | **Unit** | **Evidence class (v7)** |
| --- | --- | --- | --- | --- |
| SPC | Slides per case = slides per glass request (one case-level request); cases/yr = slides/yr / SPC | 3 | slides/case | Assumption |
| PROD | Productive share of paid hours | 0.85 | fraction | Assumption |
| FILE_MIN | On-site filing per slide (one-time, rule-invariant) | 0.25 | min/slide | Assumption |
| DESTR_MIN | On-site culling per slide at end of retention (rule-invariant at steady state) | 0.05 | min/slide | Assumption |
| PULL_MIN | Locate, pull and check out a case | 8 | min/request | Assumption |
| REFILE_MIN | Re-file returned slides | 4 | min/request | Assumption |
| TRACK_MIN | Requisition logging and tracking | 3 | min/request | Assumption |
| BLOCK_PULL | 1 = a recut request also pays one block pull, costed like a glass pull (dossier brief 5.6: 'the cost of pulling a glass slide and pulling a block are similar'); 0 = comparator convention (recut replaces the pull) | 1 | 0/1 | Assumption |
| BLDG_LIFE | Building life for annualizing construction | 40 | years | Assumption |
| FILL | Usable fill of cabinet capacity | 0.85 | fraction | Assumption |
| AISLE_W | Aisle width | 1.1 | m | Assumption |
| CIRC | Circulation allowance | 0.15 | fraction | Assumption |
| LANDED | Landed-cost uplift (freight, brokerage, tax) | 0.25 | fraction | Assumption |
| CAB_LIFE | Cabinet life for annualizing | 20 | years | Assumption |
| INTAKE_TRIPS | Transfer pick-ups per year | 12 | trips/yr | Assumption |
| BOX_PER_CARTON | Slide boxes per carton | 2 | boxes | Assumption |
| OFF_FILL | Usable fill of off-site boxes | 0.85 | fraction | Assumption |
| EV_PER_TRIP | Requests sharing one delivery and pick-up trip | 20 | requests/trip | Assumption |
| OFF_INHOUSE_MIN | In-house labour per off-site request | 6 | min/request | Assumption |
| P0_RATE | Glass requests per year per 100 cases accessioned per year, cases aged 0 to 9 y, no digital archive (LabPON about 2.7, MSKCC 2014 about 12) | 5 | requests per 100 cases/yr | Assumption (literature-informed) |
| A1_SHARE | Share of age 0 to 9 demand falling in the first 12 months; rest spread evenly over ages 1 to 9 | 0.85 | share | Assumption (literature-informed) |
| P100_RATE | Residual glass requests when the case has a retained WSI (floor; about 90% fall, MSK 93%) | 0.5 | per 100 cases/yr | Assumption (literature-informed) |
| RECUT_SHARE | BASE recut share: share of one year's slides (equivalently cases, SPC slides per request) that are requested once while aged 10 to 19 y. Steady-state recut requests per year = RECUT_SHARE x cases/yr. Per case-year request rate at ages 10 to 19 = RECUT_SHARE / 10. v6 base 0.5% (0.05% per case-yr), anchored to Ali 2023; v5 used 0.01%. No pathology source measures requests for 10 to 20 y glass | 0.005 | share of a year's slides | Illustrative assumption (no data) |
| SEL_SHARE | Share of each year's slides whose WSI is retained (selected at sign-out) | 0.1 | share of slides | Study specification (hypothetical) |
| SEL_CAPTURE | Share of digitally servable requests that fall on the retained subset (coverage). Requests on covered cases still need glass at the floor share PHI, as in S2 | 0.9 | share of requests | Study specification (hypothetical) |
| GB_40X | WSI size at 40x | 1.5 | GB/slide | Assumption (literature-informed) |
| EGRESS_SEL | Egress of each restored case once to the hospital: 1 internet egress tiers (base, conservative); 2 dedicated link (DTO only, port and circuit fees excluded); 0 none (viewer in the same cloud region) | 1 | 0/1/2 | Analyst choice |
| RESTORE_SEL | Restore volume: 1 = request-driven (each digitally served request restores its case, SPC x GB); 2 = prior model S1 convention (RECALL_D of the archive per year) | 1 | 1/2 | Analyst choice |

The recut share (RECUT_SHARE) extrapolates Ali et al. 2023 (cases aged 1 to about 4.5 years) to ages 10 to 19; no pathology study measures it, so Table S6 reports values from 0% to 2% and the break even share. The selective WSI rule (SEL_SHARE and SEL_CAPTURE) is also an assumption; its effect is shown in Table S5 and Supplementary Figure S2.

**Table S4. Steady state annual cost by laboratory, scenario, retention rule and component (CAD per year; Supplementary File 1, sheets Summary and Results_3x3)**

| **Scenario** | **Rule** | **Glass storage** | **Filing and destruction** | **Glass retrieval** | **Recuts** | **WSI archive storage** | **Restore fees** | **6 month hot copy** | **Egress** | **Total** |
| --- | --- | --- | --- | --- | --- | --- | --- | --- | --- | --- |
| **Small community laboratory** | | | | | | | | | | |
| Glass only | 10 y | 7,417 | 22,192 | 18,516 | 13,329 | 0 | 0 | 0 | 0 | 61,454 |
| Glass only | 20 y | 14,834 | 22,192 | 20,368 | 0 | 0 | 0 | 0 | 0 | 57,394 |
| Glass + WSI of every slide | 10 y | 7,417 | 22,192 | 1,852 | 13,329 | 38,117 | 215 | 1,155 | 837 | 85,114 |
| Glass + WSI of every slide | 20 y | 14,834 | 22,192 | 2,037 | -0 | 73,647 | 237 | 1,270 | 921 | 115,138 |
| Glass + selective WSI | 10 y | 7,417 | 22,192 | 3,518 | 13,329 | 5,172 | 194 | 1,039 | 754 | 53,614 |
| Glass + selective WSI | 20 y | 14,834 | 22,192 | 3,870 | 0 | 7,757 | 213 | 1,143 | 829 | 50,838 |
| **Academic laboratory** | | | | | | | | | | |
| Glass only | 10 y | 36,714 | 110,960 | 92,581 | 66,643 | 0 | 0 | 0 | 0 | 306,897 |
| Glass only | 20 y | 73,057 | 110,960 | 101,839 | 0 | 0 | 0 | 0 | 0 | 285,856 |
| Glass + WSI of every slide | 10 y | 36,714 | 110,960 | 9,258 | 66,643 | 181,488 | 1,076 | 5,774 | 4,186 | 416,098 |
| Glass + WSI of every slide | 20 y | 73,057 | 110,960 | 10,184 | -0 | 362,967 | 1,184 | 6,351 | 4,605 | 569,308 |
| Glass + selective WSI | 10 y | 36,714 | 110,960 | 17,590 | 66,643 | 20,687 | 969 | 5,197 | 3,768 | 262,526 |
| Glass + selective WSI | 20 y | 73,057 | 110,960 | 19,349 | 0 | 38,116 | 1,066 | 5,716 | 4,144 | 252,409 |
| **Large academic laboratory** | | | | | | | | | | |
| Glass only | 10 y | 54,885 | 166,440 | 138,872 | 99,964 | 0 | 0 | 0 | 0 | 460,160 |
| Glass only | 20 y | 109,399 | 166,440 | 152,759 | 0 | 0 | 0 | 0 | 0 | 428,598 |
| Glass + WSI of every slide | 10 y | 54,885 | 166,440 | 13,887 | 99,964 | 272,567 | 1,615 | 8,661 | 6,279 | 624,298 |
| Glass + WSI of every slide | 20 y | 109,399 | 166,440 | 15,276 | 0 | 541,865 | 1,776 | 9,527 | 6,907 | 851,191 |
| Glass + selective WSI | 10 y | 54,885 | 166,440 | 26,386 | 99,964 | 27,774 | 1,453 | 7,795 | 5,651 | 390,348 |
| Glass + selective WSI | 20 y | 109,399 | 166,440 | 29,024 | 0 | 55,546 | 1,598 | 8,575 | 6,217 | 376,799 |

Glass storage = floor space and cabinets (on site). Glass retrieval = labour to pull, refile and log requested cases. Recuts = block pull, recut and H&E for requests on glass discarded after 10 years. Restore fees = archive rehydration and request charges; hot copy = 6 months in the hot tier; egress = one internet download per restored case. Components sum to the total.

**Table S5. Selective WSI: cost relative to glass only under alternative selection rules (Supplementary File 1, sheets Sensitivity and Summary)**

| **Selection rule** | **Small: S3 minus S1, 10 y (CAD/yr)** | **% of S1** | **Academic (CAD/yr)** | **% of S1** | **Large (CAD/yr)** | **% of S1** | **S3 20 y vs 10 y, academic** |
| --- | --- | --- | --- | --- | --- | --- | --- |
| Base: 10% of slides cover 90% of digitally servable requests (assumption) | -7,840 | -12.8% | -44,371 | -14.5% | -69,813 | -15.2% | -3.9% |
| 5% of slides cover 80% | -8,980 | -14.6% | -47,486 | -15.5% | -71,229 | -15.5% | -6.4% |
| 20% of slides cover 95% | -5,977 | -9.7% | -30,555 | -10.0% | -47,461 | -10.3% | +2.8% |
| 10% of slides, good prediction (literature-calibrated curve, g = 7.5) | -2,726 | -4.4% | -18,800 | -6.1% | -31,456 | -6.8% | -2.6% |
| 10% of slides, imperfect prediction (literature-calibrated curve, g = 3) | +1,254 | +2.0% | +1,097 | +0.4% | -1,611 | -0.4% | -1.8% |
| 10% of slides chosen at random | +3,726 | +6.1% | +13,458 | +4.4% | +16,931 | +3.7% | -1.4% |
| 10% of slides serve 90% of all requests (no residual glass need) | -9,286 | -15.1% | -51,600 | -16.8% | -80,656 | -17.5% | -4.2% |
| Break even coverage at 10 y / 20 y (share of requests on the selected 10% of slides) | 36% | 49% | 29% | 48% | 26% | 47% |  |

Negative values mean glass plus selective WSI is cheaper than glass only under a 10 year rule. The base rule implies a capture exponent of about 21.9 on the literature-calibrated curve C(r) = 1 - (1 - r)^g, against dossier calibrations of 3 (base) and 7.5 (better prediction). In the break even row the paired columns give coverage at the 10 and 20 year rules.

**Table S6. One way sensitivity analysis by laboratory (Supplementary File 1, sheet Sensitivity)**

| **Case** | **Category** | **S1 20 y vs 10 y** | **S2 20 y vs 10 y** | **S3 20 y vs 10 y** | **S2 minus S1, 10 y (CAD/yr)** | **S3 minus S1, 10 y (CAD/yr)** |
| --- | --- | --- | --- | --- | --- | --- |
| **Small community laboratory** | | | | | | |
| Base case | base | -6.6% | +35.3% | -5.2% | +23,660 | -7,840 |
| Recut share 0 | recut share | +15.4% | +59.8% | +24.8% | +23,660 | -7,840 |
| Recut share 0.001% | recut share | +15.4% | +59.8% | +24.7% | +23,660 | -7,840 |
| Recut share 0.1% (dossier low R_OLD x 10) | recut share | +10.1% | +54.2% | +17.3% | +23,660 | -7,840 |
| Recut share 0.01% (v5 base, author proposal) | recut share | +14.9% | +59.2% | +24.0% | +23,660 | -7,840 |
| Recut share 1% | recut share | -20.8% | +17.4% | -23.2% | +23,660 | -7,840 |
| Recut share = prior model S3 flat tail (0.83%) | recut share | -16.6% | +22.8% | -18.1% | +23,660 | -7,840 |
| Recut share = prior model S2 OLD_RECALL x 10 (2%) | recut share | -37.9% | -7.0% | -43.9% | +23,660 | -7,840 |
| Requests for discarded glass not costed (dossier convention) | recut share | +19.3% | +60.4% | +26.2% | +23,660 | -7,840 |
| Recut replaces the pull (no block pull; comparator convention) | recut share | -3.7% | +38.3% | -1.8% | +23,660 | -7,840 |
| Selective WSI 5% of slides cover 80% of requests | coverage | -6.6% | +35.3% | -5.0% | +23,660 | -8,980 |
| Selective WSI 20% of slides cover 95% of requests | coverage | -6.6% | +35.3% | +4.2% | +23,660 | -5,977 |
| Selective WSI random: 10% of slides cover 10% of requests | coverage | -6.6% | +35.3% | -2.5% | +23,660 | +3,726 |
| Selective WSI 10%, literature-calibrated base capture curve (g = 3) | coverage | -6.6% | +35.3% | -3.0% | +23,660 | +1,254 |
| Selective WSI 10%, literature-calibrated better-prediction curve (g = 7.5) | coverage | -6.6% | +35.3% | -3.9% | +23,660 | -2,726 |
| Selective WSI: study specification literal, 90% of all requests served digitally (no residual glass floor in S3) | coverage | -6.6% | +35.3% | -5.6% | +23,660 | -9,286 |
| Tier: AWS Glacier Deep Archive (list) | tier | -6.6% | +43.6% | -1.4% | +31,895 | -8,256 |
| Tier: Azure Archive LRS pay-as-you-go | tier | -6.6% | +43.7% | -1.4% | +31,737 | -8,392 |
| Tier: Google Cloud Archive | tier | -6.6% | +52.5% | +2.0% | +50,463 | -5,914 |
| Scan resolution 20x (0.5 GB/slide) | resolution | -6.6% | +11.3% | -11.1% | -2,996 | -11,748 |
| Request rate 2.5 per 100 cases | requests | -7.8% | +36.1% | -5.5% | +30,889 | -1,334 |
| Request rate 12 per 100 cases | requests | -4.6% | +33.1% | -4.5% | +3,420 | -26,056 |
| Residual glass share low (0.2 per 100) | requests | -6.6% | +35.6% | -5.4% | +22,696 | -8,707 |
| Residual glass share high (1.7 per 100) | requests | -6.6% | +34.2% | -4.3% | +27,515 | -4,370 |
| Glass retrieval 10 min per request | glass | -7.4% | +36.5% | -4.4% | +29,208 | -2,847 |
| Glass retrieval 20 min per request | glass | -6.0% | +34.1% | -5.9% | +18,112 | -12,833 |
| Glass on site, off-site retrieval cost per request | glass | -5.0% | +31.9% | -7.3% | +6,928 | -22,898 |
| Glass archive off site (vendor storage, intake, fees, retrieval) | glass | +1.3% | +8.6% | +1.3% | +6,928 | -22,898 |
| Glass storage continuous per slide-year (no whole stacks) | glass | -6.9% | +35.2% | -5.5% | +23,660 | -7,840 |
| Discount 0% (cabinet annualization) | discount | -7.7% | +34.8% | -6.5% | +23,660 | -7,840 |
| Discount 3% (cabinet annualization) | discount | -5.4% | +35.8% | -3.8% | +23,660 | -7,840 |
| Hot hold of restores 3 months | digital | -6.6% | +35.4% | -5.3% | +23,082 | -8,360 |
| Hot hold of restores 12 months | digital | -6.6% | +34.9% | -4.9% | +24,815 | -6,801 |
| No egress (viewer in the cloud region) | digital | -6.6% | +35.5% | -5.4% | +22,823 | -8,593 |
| Egress over a dedicated link (DTO only) | digital | -6.6% | +35.5% | -5.3% | +23,063 | -8,377 |
| Restore volume = dossier convention, 5% of archive per year | digital | -6.6% | +50.1% | -1.0% | +45,979 | -7,374 |
| **Academic laboratory** | | | | | | |
| Base case | base | -6.9% | +36.8% | -3.9% | +109,201 | -44,371 |
| Recut share 0 | recut share | +15.1% | +62.3% | +27.5% | +109,201 | -44,371 |
| Recut share 0.001% | recut share | +15.1% | +62.3% | +27.4% | +109,201 | -44,371 |
| Recut share 0.1% (dossier low R_OLD x 10) | recut share | +9.8% | +56.5% | +19.6% | +109,201 | -44,371 |
| Recut share 0.01% (v5 base, author proposal) | recut share | +14.6% | +61.7% | +26.6% | +109,201 | -44,371 |
| Recut share 1% | recut share | -21.0% | +18.4% | -22.5% | +109,201 | -44,371 |
| Recut share = prior model S3 flat tail (0.83%) | recut share | -16.9% | +23.9% | -17.2% | +109,201 | -44,371 |
| Recut share = prior model S2 OLD_RECALL x 10 (2%) | recut share | -38.1% | -6.6% | -43.6% | +109,201 | -44,371 |
| Requests for discarded glass not costed (dossier convention) | recut share | +19.0% | +62.9% | +28.9% | +109,201 | -44,371 |
| Recut replaces the pull (no block pull; comparator convention) | recut share | -4.0% | +39.9% | -0.3% | +109,201 | -44,371 |
| Selective WSI 5% of slides cover 80% of requests | coverage | -6.9% | +36.8% | -6.4% | +109,201 | -47,486 |
| Selective WSI 20% of slides cover 95% of requests | coverage | -6.9% | +36.8% | +2.8% | +109,201 | -30,555 |
| Selective WSI random: 10% of slides cover 10% of requests | coverage | -6.9% | +36.8% | -1.4% | +109,201 | +13,458 |
| Selective WSI 10%, literature-calibrated base capture curve (g = 3) | coverage | -6.9% | +36.8% | -1.8% | +109,201 | +1,097 |
| Selective WSI 10%, literature-calibrated better-prediction curve (g = 7.5) | coverage | -6.9% | +36.8% | -2.6% | +109,201 | -18,800 |
| Selective WSI: study specification literal, 90% of all requests served digitally (no residual glass floor in S3) | coverage | -6.9% | +36.8% | -4.2% | +109,201 | -51,600 |
| Tier: AWS Glacier Deep Archive (list) | tier | -6.9% | +43.5% | -1.7% | +159,473 | -41,279 |
| Tier: Azure Archive LRS pay-as-you-go | tier | -6.9% | +43.5% | -1.7% | +158,685 | -41,961 |
| Tier: Google Cloud Archive | tier | -6.9% | +52.4% | +1.8% | +252,317 | -29,569 |
| Scan resolution 20x (0.5 GB/slide) | resolution | -6.9% | +11.0% | -9.5% | -18,908 | -63,911 |
| Request rate 2.5 per 100 cases | requests | -8.1% | +37.7% | -4.1% | +145,344 | -11,842 |
| Request rate 12 per 100 cases | requests | -4.8% | +34.5% | -3.4% | +8,000 | -135,452 |
| Residual glass share low (0.2 per 100) | requests | -6.9% | +37.1% | -4.1% | +104,382 | -48,708 |
| Residual glass share high (1.7 per 100) | requests | -6.9% | +35.6% | -3.0% | +128,478 | -27,022 |
| Glass retrieval 10 min per request | glass | -7.7% | +38.1% | -3.0% | +136,941 | -19,405 |
| Glass retrieval 20 min per request | glass | -6.2% | +35.6% | -4.6% | +81,461 | -69,337 |
| Glass on site, off-site retrieval cost per request | glass | -5.1% | +33.3% | -6.1% | +25,543 | -119,663 |
| Glass archive off site (vendor storage, intake, fees, retrieval) | glass | +1.3% | +8.9% | +1.6% | +25,543 | -119,663 |
| Glass storage continuous per slide-year (no whole stacks) | glass | -6.9% | +36.9% | -3.9% | +109,201 | -44,371 |
| Discount 0% (cabinet annualization) | discount | -8.0% | +36.3% | -5.1% | +109,201 | -44,371 |
| Discount 3% (cabinet annualization) | discount | -5.7% | +37.3% | -2.5% | +109,201 | -44,371 |
| Hot hold of restores 3 months | digital | -6.9% | +37.0% | -4.0% | +106,314 | -46,969 |
| Hot hold of restores 12 months | digital | -6.9% | +36.5% | -3.6% | +114,975 | -39,174 |
| No egress (viewer in the cloud region) | digital | -6.9% | +37.1% | -4.1% | +105,015 | -48,139 |
| Egress over a dedicated link (DTO only) | digital | -6.9% | +37.0% | -4.0% | +106,218 | -47,056 |
| Restore volume = dossier convention, 5% of archive per year | digital | -6.9% | +51.0% | +0.4% | +217,498 | -42,041 |
| **Large academic laboratory** | | | | | | |
| Base case | base | -6.9% | +36.3% | -3.5% | +164,137 | -69,813 |
| Recut share 0 | recut share | +15.1% | +61.8% | +28.3% | +164,137 | -69,813 |
| Recut share 0.001% | recut share | +15.1% | +61.7% | +28.3% | +164,137 | -69,813 |
| Recut share 0.1% (dossier low R_OLD x 10) | recut share | +9.8% | +55.9% | +20.3% | +164,137 | -69,813 |
| Recut share 0.01% (v5 base, author proposal) | recut share | +14.6% | +61.2% | +27.5% | +164,137 | -69,813 |
| Recut share 1% | recut share | -21.0% | +17.9% | -22.3% | +164,137 | -69,813 |
| Recut share = prior model S3 flat tail (0.83%) | recut share | -16.9% | +23.5% | -16.9% | +164,137 | -69,813 |
| Recut share = prior model S2 OLD_RECALL x 10 (2%) | recut share | -38.1% | -6.9% | -43.6% | +164,137 | -69,813 |
| Requests for discarded glass not costed (dossier convention) | recut share | +19.0% | +62.3% | +29.8% | +164,137 | -69,813 |
| Recut replaces the pull (no block pull; comparator convention) | recut share | -4.0% | +39.4% | +0.1% | +164,137 | -69,813 |
| Selective WSI 5% of slides cover 80% of requests | coverage | -6.9% | +36.3% | -7.2% | +164,137 | -71,229 |
| Selective WSI 20% of slides cover 95% of requests | coverage | -6.9% | +36.3% | +2.7% | +164,137 | -47,461 |
| Selective WSI random: 10% of slides cover 10% of requests | coverage | -6.9% | +36.3% | -1.0% | +164,137 | +16,931 |
| Selective WSI 10%, literature-calibrated base capture curve (g = 3) | coverage | -6.9% | +36.3% | -1.5% | +164,137 | -1,611 |
| Selective WSI 10%, literature-calibrated better-prediction curve (g = 7.5) | coverage | -6.9% | +36.3% | -2.3% | +164,137 | -31,456 |
| Selective WSI: study specification literal, 90% of all requests served digitally (no residual glass floor in S3) | coverage | -6.9% | +36.3% | -3.9% | +164,137 | -80,656 |
| Tier: AWS Glacier Deep Archive (list) | tier | -6.9% | +43.5% | -1.7% | +239,209 | -61,919 |
| Tier: Azure Archive LRS pay-as-you-go | tier | -6.9% | +43.5% | -1.7% | +238,027 | -62,941 |
| Tier: Google Cloud Archive | tier | -6.9% | +52.4% | +1.8% | +378,475 | -44,354 |
| Scan resolution 20x (0.5 GB/slide) | resolution | -6.9% | +10.9% | -8.8% | -28,362 | -97,159 |
| Request rate 2.5 per 100 cases | requests | -8.1% | +37.3% | -3.7% | +218,352 | -21,020 |
| Request rate 12 per 100 cases | requests | -4.8% | +34.0% | -3.0% | +12,205 | -206,482 |
| Residual glass share low (0.2 per 100) | requests | -6.9% | +36.7% | -3.7% | +156,908 | -76,319 |
| Residual glass share high (1.7 per 100) | requests | -6.9% | +35.2% | -2.6% | +193,052 | -43,790 |
| Glass retrieval 10 min per request | glass | -7.7% | +37.6% | -2.6% | +205,747 | -32,364 |
| Glass retrieval 20 min per request | glass | -6.2% | +35.2% | -4.3% | +122,527 | -107,262 |
| Glass on site, off-site retrieval cost per request | glass | -5.1% | +32.9% | -5.8% | +38,650 | -182,751 |
| Glass archive off site (vendor storage, intake, fees, retrieval) | glass | +1.3% | +8.8% | +1.6% | +38,650 | -182,751 |
| Glass storage continuous per slide-year (no whole stacks) | glass | -6.9% | +36.4% | -3.5% | +164,137 | -69,813 |
| Discount 0% (cabinet annualization) | discount | -8.0% | +35.9% | -4.8% | +164,137 | -69,813 |
| Discount 3% (cabinet annualization) | discount | -5.7% | +36.9% | -2.1% | +164,137 | -69,813 |
| Hot hold of restores 3 months | digital | -6.9% | +36.5% | -3.6% | +159,806 | -73,710 |
| Hot hold of restores 12 months | digital | -6.9% | +36.0% | -3.2% | +172,798 | -62,018 |
| No egress (viewer in the cloud region) | digital | -6.9% | +36.6% | -3.7% | +157,858 | -75,464 |
| Egress over a dedicated link (DTO only) | digital | -6.9% | +36.5% | -3.6% | +159,662 | -73,840 |
| Restore volume = dossier convention, 5% of archive per year | digital | -6.9% | +49.9% | +0.8% | +325,878 | -66,318 |

Percentages are changes in steady state annual cost from a 20 year over a 10 year rule (negative = 20 year rule cheaper). The last two columns compare scenarios under the 10 year rule (negative = digital scenario cheaper). The discount rate affects only the annualization of cabinets in the steady state outcome. 'Glass archive off site' replaces on site storage and retrieval by vendor tariffs (Iron Mountain pathology lines, a US public sector tariff used as a proxy for Canadian rates; see Table S10 for the full off-site analysis). Recut share rows: share of one year's slides requested at ages 10 to 19 years (base 0.5%); the glass only 20 year rule costs more than the 10 year rule only below about 0.32%. Cloud tier, 20x scanning and off-site storage change several linked costs and are scenarios rather than one-way parameter changes.

**Table S7. Steady state annual cost under the 8, 10 and 20 year jurisdiction rules (CAD per year; Supplementary File 1, sheets Jurisdictions and Summary)**

| **Rule** | **Scenario** | **Small community** | **Academic** | **Large academic** |
| --- | --- | --- | --- | --- |
| UK RCPath 8 y | Scenario 1 glass only | 63,796 | 318,609 | 477,727 |
| UK RCPath 8 y | Scenario 2 glass + WSI of every slide | 80,180 | 395,360 | 589,933 |
| UK RCPath 8 y | Scenario 3 glass + selective WSI (10% of slides, 90% coverage) | 56,390 | 271,235 | 406,666 |
| US CLIA/CAP 10 y | Scenario 1 glass only | 61,454 | 306,897 | 460,160 |
| US CLIA/CAP 10 y | Scenario 2 glass + WSI of every slide | 85,114 | 416,098 | 624,298 |
| US CLIA/CAP 10 y | Scenario 3 glass + selective WSI (10% of slides, 90% coverage) | 53,614 | 262,526 | 390,348 |
| CAP-ACP 2005; BC 2024 20 y | Scenario 1 glass only | 57,394 | 285,856 | 428,598 |
| CAP-ACP 2005; BC 2024 20 y | Scenario 2 glass + WSI of every slide | 115,138 | 569,308 | 851,191 |
| CAP-ACP 2005; BC 2024 20 y | Scenario 3 glass + selective WSI (10% of slides, 90% coverage) | 50,838 | 252,409 | 376,799 |
| **Change relative to the 10 year rule** | | | | |
| 8 y (UK) vs 10 y | Glass only | +3.8% | +3.8% | +3.8% |
| 8 y (UK) vs 10 y | Glass + WSI of every slide | -5.8% | -5.0% | -5.5% |
| 8 y (UK) vs 10 y | Glass + selective WSI | +5.2% | +3.3% | +4.2% |
| 20 y (Canada, BC) vs 10 y | Glass only | -6.6% | -6.9% | -6.9% |
| 20 y (Canada, BC) vs 10 y | Glass + WSI of every slide | +35.3% | +36.8% | +36.3% |
| 20 y (Canada, BC) vs 10 y | Glass + selective WSI | -5.2% | -3.9% | -3.5% |

Rule labels give the source of each period. Within Canada the rules conflict: the Ontario regulation requires at least 10 years, while the CAP-ACP and British Columbia guidance recommend 20 years. The 8 year glass only rule costs more than the 10 year rule because requests at ages 8 and 9 years carry the age 1 to 9 request rate and must be recut once glass is discarded. Glass and images are kept for the same period in every scenario. Plotted in Supplementary Figure S1.

**Table S8. Mixed retention rules: different periods for glass and images (steady state CAD per year; Supplementary File 1, sheet Summary, MIX_*)**

| **Scenario** | **Rule** | **Small community** | **Academic** | **Large academic** |
| --- | --- | --- | --- | --- |
| Glass + WSI of every slide | Glass 10 y, WSI 10 y | 85,114 | 416,098 | 624,298 |
| Glass + WSI of every slide | Glass 20 y, WSI 10 y | 81,054 | 395,057 | 592,735 |
| Glass + WSI of every slide | Glass 10 y, WSI 20 y | 108,868 | 538,704 | 805,284 |
| Glass + WSI of every slide | Glass 20 y, WSI 20 y | 115,138 | 569,308 | 851,191 |
| Glass + selective WSI | Glass 10 y, WSI 10 y | 53,614 | 262,526 | 390,348 |
| Glass + selective WSI | Glass 20 y, WSI 10 y | 49,554 | 241,485 | 358,785 |
| Glass + selective WSI | Glass 10 y, WSI 20 y | 45,602 | 226,969 | 338,640 |
| Glass + selective WSI | Glass 20 y, WSI 20 y | 50,838 | 252,409 | 376,799 |

Medium specific rules: keeping glass for 20 years and images for 10 years costs much less than keeping both for 20 years when every slide is imaged.

**Table S9. Secondary outcome: net present cost over 30 years from an empty archive (CAD; Supplementary File 1, sheets Calc_NPC and Summary)**

| **Scenario** | **NPC, 10 y rule** | **NPC, 20 y rule** | **20 y minus 10 y** | **Change (1.5%)** | **Change (0%)** | **Change (3%)** | **Undiscounted, 10 y rule** | **Undiscounted, 20 y rule** |
| --- | --- | --- | --- | --- | --- | --- | --- | --- |
| **Small community laboratory** | | | | | | | | |
| Glass only | 1,243,471 | 1,167,814 | -75,657 | -6.1% | -6.4% | -5.8% | 1,566,095 | 1,466,179 |
| Glass + WSI of every slide | 1,670,477 | 1,978,672 | +308,195 | +18.4% | +20.2% | +16.7% | 2,120,100 | 2,547,482 |
| Glass + selective WSI | 1,046,796 | 988,693 | -58,102 | -5.6% | -5.7% | -5.3% | 1,325,142 | 1,249,016 |
| **Academic laboratory** | | | | | | | | |
| Glass only | 6,210,114 | 5,822,244 | -387,870 | -6.2% | -6.6% | -5.9% | 7,821,206 | 7,308,646 |
| Glass + WSI of every slide | 8,148,506 | 9,726,705 | +1,578,199 | +19.4% | +21.1% | +17.6% | 10,344,901 | 12,532,323 |
| Glass + selective WSI | 5,088,512 | 4,819,529 | -268,983 | -5.3% | -5.4% | -5.1% | 6,448,367 | 6,099,572 |
| **Large academic laboratory** | | | | | | | | |
| Glass only | 9,311,191 | 8,729,851 | -581,340 | -6.2% | -6.6% | -5.9% | 11,726,803 | 10,958,519 |
| Glass + WSI of every slide | 12,213,598 | 14,547,039 | +2,333,441 | +19.1% | +20.9% | +17.4% | 15,508,324 | 18,743,133 |
| Glass + selective WSI | 7,573,012 | 7,201,583 | -371,429 | -4.9% | -5.0% | -4.8% | 9,594,613 | 9,113,686 |

NPC = net present cost at a 1.5% real discount rate (0% and 3% in sensitivity), 30 year horizon starting from an empty archive, so a 20 year rule runs for 10 years at steady state. Cabinets are charged as an equivalent annual cost while in use (stacks in use x landed cost x capital recovery factor), consistent with the steady state model; reserved cloud capacity is charged at one third of each 3 year commitment per year.

**Table S10. Secondary analysis: glass slides kept on site or with a commercial off-site vendor (steady state CAD per year; in brackets, change versus all on site; Supplementary File 1, sheet OffSite)**

| **Scenario** | **Glass storage option, rule** | **Small community** | **Academic** | **Large academic** |
| --- | --- | --- | --- | --- |
| Glass only | All on site (base), 10 y | 61,454 | 306,897 | 460,160 |
| Glass only | All on site (base), 20 y | 57,394 | 285,856 | 428,598 |
| Glass only | On site, space at new hospital build cost, 10 y | 64,748 (+5.4%) | 323,207 (+5.3%) | 484,542 (+5.3%) |
| Glass only | On site, space at new hospital build cost, 20 y | 63,983 (+11.5%) | 318,310 (+11.4%) | 477,197 (+11.3%) |
| Glass only | 1 y on site, then vendor, records lines, 10 y | 89,269 (+45.3%) | 439,715 (+43.3%) | 658,744 (+43.2%) |
| Glass only | 1 y on site, then vendor, records lines, 20 y | 92,972 (+62.0%) | 458,234 (+60.3%) | 686,522 (+60.2%) |
| Glass only | 1 y on site, then vendor, pathology lines, 10 y | 436,536 (+610.4%) | 2,176,051 (+609.0%) | 3,263,247 (+609.2%) |
| Glass only | 1 y on site, then vendor, pathology lines, 20 y | 442,316 (+670.7%) | 2,204,949 (+671.4%) | 3,306,595 (+671.5%) |
| Glass only | Vendor from accession, records lines, 10 y | 90,018 (+46.5%) | 443,463 (+44.5%) | 664,365 (+44.4%) |
| Glass only | Vendor from accession, records lines, 20 y | 93,722 (+63.3%) | 461,981 (+61.6%) | 692,143 (+61.5%) |
| Glass only | Vendor from accession, pathology lines, 10 y | 434,643 (+607.3%) | 2,166,585 (+606.0%) | 3,249,049 (+606.1%) |
| Glass only | Vendor from accession, pathology lines, 20 y | 440,422 (+667.4%) | 2,195,484 (+668.0%) | 3,292,397 (+668.2%) |
| Glass + WSI of every slide | All on site (base), 10 y | 85,114 | 416,098 | 624,298 |
| Glass + WSI of every slide | All on site (base), 20 y | 115,138 | 569,308 | 851,191 |
| Glass + WSI of every slide | On site, space at new hospital build cost, 10 y | 88,408 (+3.9%) | 432,408 (+3.9%) | 648,679 (+3.9%) |
| Glass + WSI of every slide | On site, space at new hospital build cost, 20 y | 121,727 (+5.7%) | 601,762 (+5.7%) | 899,789 (+5.7%) |
| Glass + WSI of every slide | 1 y on site, then vendor, records lines, 10 y | 109,961 (+29.2%) | 534,077 (+28.4%) | 800,622 (+28.2%) |
| Glass + WSI of every slide | 1 y on site, then vendor, records lines, 20 y | 145,770 (+26.6%) | 716,954 (+25.9%) | 1,072,017 (+25.9%) |
| Glass + WSI of every slide | 1 y on site, then vendor, pathology lines, 10 y | 457,686 (+437.7%) | 2,272,703 (+446.2%) | 3,408,561 (+446.0%) |
| Glass + WSI of every slide | 1 y on site, then vendor, pathology lines, 20 y | 495,877 (+330.7%) | 2,467,487 (+333.4%) | 3,697,816 (+334.4%) |
| Glass + WSI of every slide | Vendor from accession, records lines, 10 y | 93,893 (+10.3%) | 453,736 (+9.0%) | 680,111 (+8.9%) |
| Glass + WSI of every slide | Vendor from accession, records lines, 20 y | 129,702 (+12.6%) | 636,614 (+11.8%) | 951,506 (+11.8%) |
| Glass + WSI of every slide | Vendor from accession, pathology lines, 10 y | 441,571 (+418.8%) | 2,192,128 (+426.8%) | 3,287,699 (+426.6%) |
| Glass + WSI of every slide | Vendor from accession, pathology lines, 20 y | 479,762 (+316.7%) | 2,386,912 (+319.3%) | 3,576,954 (+320.2%) |
| Glass + selective WSI | All on site (base), 10 y | 53,614 | 262,526 | 390,348 |
| Glass + selective WSI | All on site (base), 20 y | 50,838 | 252,409 | 376,799 |
| Glass + selective WSI | On site, space at new hospital build cost, 10 y | 56,909 (+6.1%) | 278,836 (+6.2%) | 414,729 (+6.2%) |
| Glass + selective WSI | On site, space at new hospital build cost, 20 y | 57,428 (+13.0%) | 284,863 (+12.9%) | 425,398 (+12.9%) |
| Glass + selective WSI | 1 y on site, then vendor, records lines, 10 y | 78,758 (+46.9%) | 381,989 (+45.5%) | 568,898 (+45.7%) |
| Glass + selective WSI | 1 y on site, then vendor, records lines, 20 y | 81,965 (+61.2%) | 402,528 (+59.5%) | 601,335 (+59.6%) |
| Glass + selective WSI | 1 y on site, then vendor, pathology lines, 10 y | 426,437 (+695.4%) | 2,120,386 (+707.7%) | 3,176,494 (+713.8%) |
| Glass + selective WSI | 1 y on site, then vendor, pathology lines, 20 y | 431,996 (+749.7%) | 2,152,679 (+752.9%) | 3,226,562 (+756.3%) |
| Glass + selective WSI | Vendor from accession, records lines, 10 y | 64,371 (+20.1%) | 310,057 (+18.1%) | 461,001 (+18.1%) |
| Glass + selective WSI | Vendor from accession, records lines, 20 y | 67,579 (+32.9%) | 330,596 (+31.0%) | 493,438 (+31.0%) |
| Glass + selective WSI | Vendor from accession, pathology lines, 10 y | 411,744 (+668.0%) | 2,046,922 (+679.7%) | 3,066,298 (+685.5%) |
| Glass + selective WSI | Vendor from accession, pathology lines, 20 y | 417,303 (+720.8%) | 2,079,215 (+723.7%) | 3,116,366 (+727.1%) |

Vendor prices: Iron Mountain public national price list (Attachment D, response to RFP #250054, Public Promise Procurement, 2025), USD converted at 1.4257. Records lines: slides sent in 1.2 cu ft cartons indexed by the laboratory (storage USD 2.15 per cu ft per month; receiving USD 1.97 per cu ft; carton retrieval and refile USD 7.04 per cu ft). Pathology lines: each case received, listed, retrieved and refiled individually (temperature controlled storage USD 2.55 per cu ft per month; receiving and listing USD 7.27 per case; retrieval and refile USD 7.02 per case). Per slide (CAD): intake 0.21 (records) and 3.67 (pathology); storage 0.0130 and 0.0154 per slide-year versus 0.0073 on site; 24.30 and 22.26 per off-site request versus 11.11 on site. Hybrid: slides filed on site, kept on site for 1 year (when most first-decade requests occur), then packed (0.25 min per slide) and sent to the vendor; the vendor destroys slides at the end of retention. Paraffin blocks stay on site in every option, so recuts always pay the on-site block pull. Vendor terms, packing density and courier batching are assumptions that would need confirmation for Canadian pathology material. New hospital build: on site floor space valued at CAD 610 instead of 282 per m2 per year. On site storage would cost as much as the 1 year hybrid (records lines) only if floor space cost about CAD 2,027 per m2 per year (academic laboratory, glass only, 20 year rule).

**Table S11. Break-even thresholds, glass only: change in steady state annual cost of a 20 year over a 10 year rule (CAD per year, academic laboratory) by second-decade request share and recut and H&E cost per slide (Supplementary File 1, sheet Threshold)**

| **Share \ CAD per recut slide** | **5** | **10** | **13.22** | **15** | **22.95** | **30** | **45** | **60** |
| --- | --- | --- | --- | --- | --- | --- | --- | --- |
| 0.00% | +36,343 | +36,343 | +36,343 | +36,343 | +36,343 | +36,343 | +36,343 | +36,343 |
| 0.05% | +35,093 | +33,843 | +33,038 | +32,593 | +30,605 | +28,843 | +25,093 | +21,343 |
| 0.10% | +33,843 | +31,343 | +29,733 | +28,843 | +24,868 | +21,343 | +13,843 | +6,343 |
| 0.20% | +31,343 | +26,343 | +23,123 | +21,343 | +13,393 | +6,343 | -8,657 | -23,657 |
| 0.29% | +29,093 | +21,843 | +17,174 | +14,593 | +3,065 | -7,157 | -28,907 | -50,657 |
| 0.50% | +23,843 | +11,343 | +3,293 | -1,157 | -21,032 | -38,657 | -76,157 | -113,657 |
| 1.00% | +11,343 | -13,657 | -29,757 | -38,657 | -78,407 | -113,657 | -188,657 | -263,657 |
| 2.00% | -13,657 | -63,657 | -95,857 | -113,657 | -193,157 | -263,657 | -413,657 | -563,657 |

Positive values: the 20 year rule costs more. No published data describe the share of slides requested at ages 10 to 19 years; the base value 0.5% is illustrative. Break even: share x recut cost per slide = second-decade storage cost per slide (CAD 0.0742 / 0.0727 / 0.0727). At the base recut cost (CAD 22.95 per slide, a US academic fee schedule used as a proxy) the break-even share is 0.323% / 0.317% / 0.317%; at the base share the break-even recut cost is CAD 14.83 / 14.54 / 14.54 per slide (small / academic / large). Selective WSI: the base-case saving over glass only under a 10 year rule would be cancelled by selection effort of CAD 0.235 / 0.266 / 0.279 per case accessioned (0.32 / 0.36 / 0.38 minutes of laboratory assistant time); with the imperfect prediction curve (g = 3) the saving per case is -0.038 / -0.007 / 0.006 CAD. Selection effort is not costed in the model. Plotted in Supplementary Figure S3.

**Supplementary Figure S1**

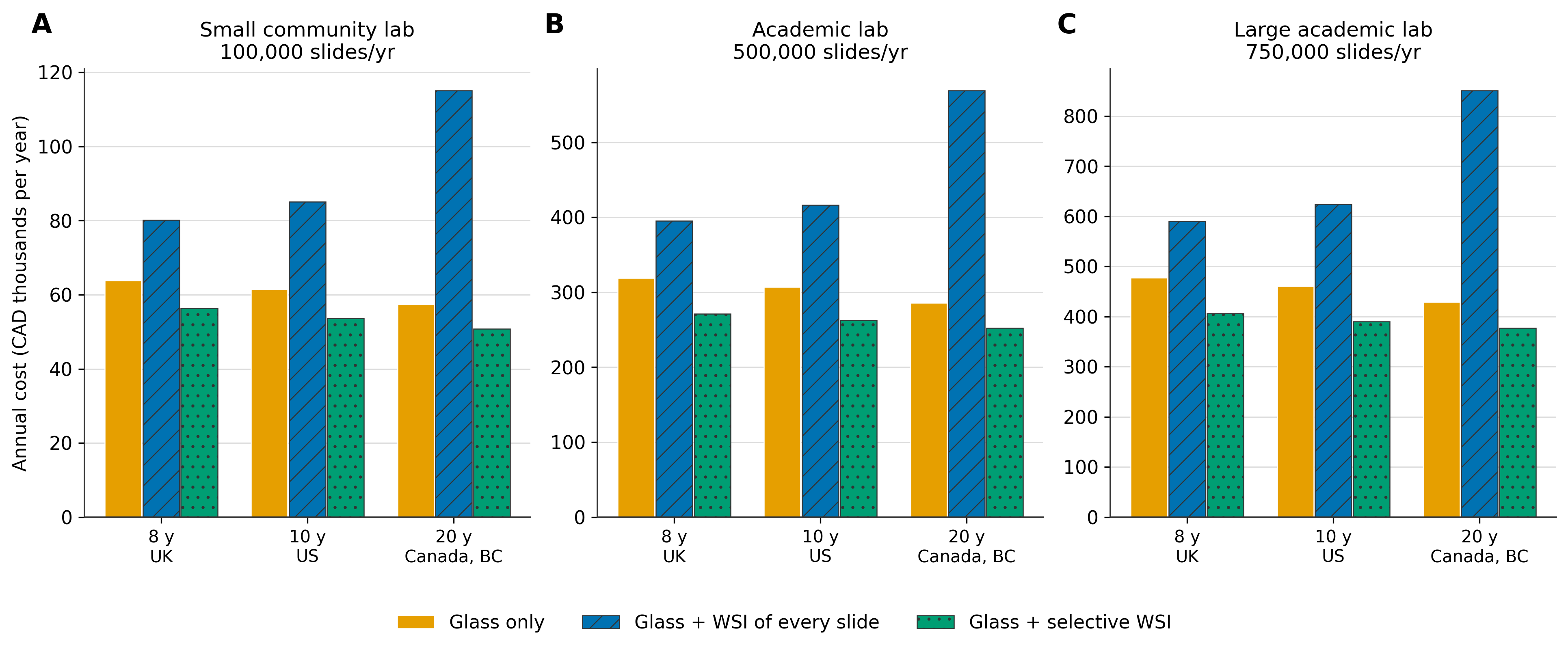

Supplementary Figure S1. Steady state annual cost implied by the minimum glass retention rules of the United Kingdom (8 years, if the slide can be recut from the block), the United States (10 years) and the Canadian Association of Pathologists and British Columbia (20 years). Within Canada the rules conflict: the Ontario regulation requires at least 10 years. (A) small community, (B) academic, (C) large academic laboratory. Relative to a 10 year rule (small / academic / large), a 20 year rule changes cost by -6.6% / -6.9% / -6.9% (glass only), +35.3% / +36.8% / +36.3% (glass plus WSI of every slide) and -5.2% / -3.9% / -3.5% (glass plus selective WSI); an 8 year rule by +3.8% / +3.8% / +3.8%, -5.8% / -5.0% / -5.5% and +5.2% / +3.3% / +4.2%. The 8 year glass only rule costs more than the 10 year rule in this model because requests at ages 8 and 9 years are frequent enough that recuts outweigh the storage saved. Values: Supplementary File 1, sheets Jurisdictions and Summary (JUR_*).

**Supplementary Figure S2**

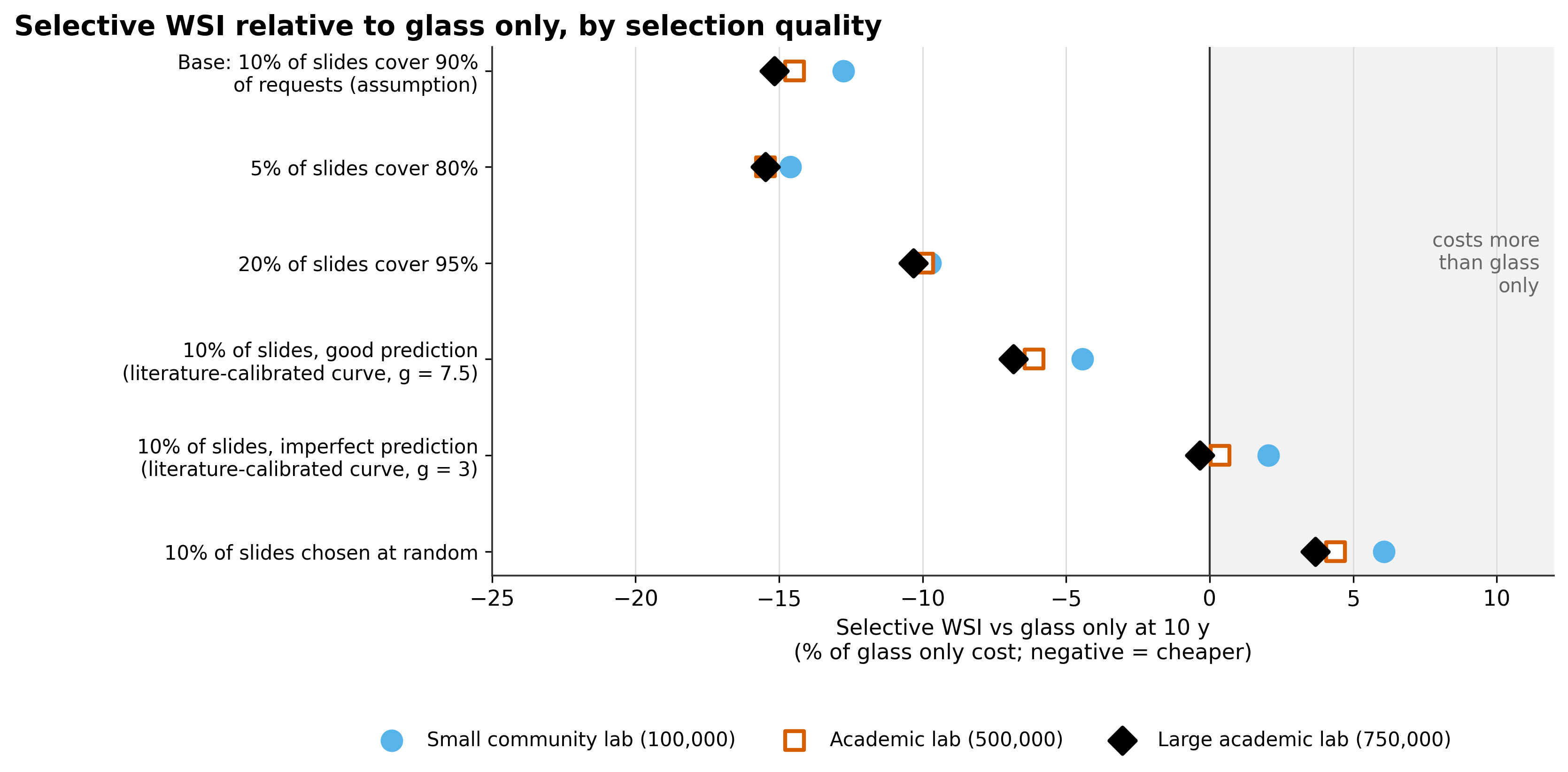

Supplementary Figure S2. Cost of glass plus selective WSI relative to glass only under a 10 year rule, for each laboratory and selection rule. g is the exponent of the capture curve calibrated in earlier work on published selection data. The base rule (10% of slides assumed to cover 90% of requests) saves 12.8% / 14.5% / 15.2% of the glass only cost; with the imperfect prediction curve (g = 3) the difference is +1,254 / +1,097 / -1,611 CAD per year (about break even) and with random selection selective WSI costs more (+3,726 / +13,458 / +16,931 CAD per year) (small / academic / large). Selective WSI breaks even with glass only when the selected slides cover about 36% / 29% / 26% of requests (10 year rule) or 49% / 48% / 47% (20 year rule). WSI, whole slide images; y, years; CAD, Canadian dollars (2026). Values: Supplementary File 1, sheets Sensitivity and Summary (SENS_cov*, SEL_*).

**Supplementary Figure S3**

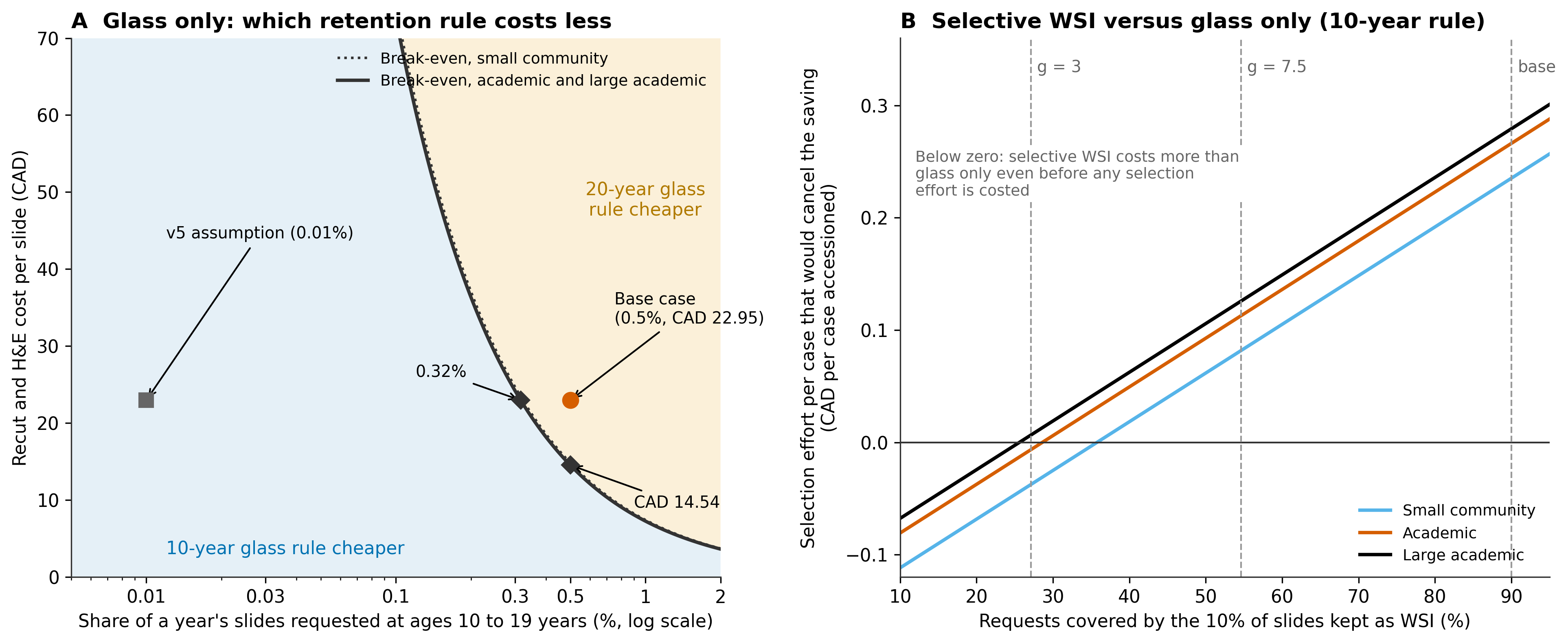

Supplementary Figure S3. Break-even thresholds. (A) Glass only: the retention rule that costs less, by second-decade request share (share of one year's slides requested at ages 10 to 19 years, log scale) and recut and H&E cost per slide. Above the line a 20 year glass rule costs less, because the recuts it avoids cost more than storing the glass for a second decade; below it a 10 year rule costs less. Base case: 0.5% (illustrative; no published data) and CAD 22.95 per slide (a US academic fee schedule used as a proxy); the square marks the version 5 assumption of 0.01%. (B) Selective WSI versus glass only under a 10 year rule: selection effort per case accessioned that would cancel the modelled saving, by the share of digitally servable requests covered by the 10% of slides kept as WSI (linear between random selection and the base rule); g = 3 and g = 7.5 mark the illustrative capture curves. Selection effort is not costed in the model. WSI, whole slide images; CAD, Canadian dollars (2026). Values: Supplementary File 1, sheet Threshold; Table S11.
